# Sex differences in functional network topology over the course of aging in 37543 UK Biobank participants

**DOI:** 10.1101/2022.03.08.22272089

**Authors:** Mite Mijalkov, Dániel Veréb, Oveis Jamialahmadi, Anna Canal-Garcia, Emiliano Gómez-Ruiz, Didac Vidal-Piñeiro, Stefano Romeo, Giovanni Volpe, Joana B. Pereira

**Author notes:** Corresponding authors: //. Address: KI, Dept. NVS, division of clinical geriatrics, Neo 7th floor, Blickagången 16, 141 83 Huddinge, Sweden.

## Abstract

Aging is a major risk factor for cardiovascular and neurodegenerative disorders, with considerable societal and economic implications. Healthy aging is accompanied by changes in connectivity between and within resting-state functional networks, however, there is no consensus on the impact of sex on these age-related alterations. Here, in a large cross-sectional sample of 37543 UK Biobank participants, we show that multilayer measures that capture the interaction between positive and negative connections provide crucial information on the impact of sex on age-related changes in network topology, being closely related to cognitive, structural, and cardiovascular risk factors that have been shown to differ between men and women. We also provide additional insights into the genetic influences on multilayer connectivity changes that occur during aging. Our findings indicate that multilayer measures contain previously unknown information on the relationship between sex and age, opening up new avenues for research into functional brain connectivity in aging.

## I. INTRODUCTION

Although human lifespan has increased from 50 to 80 years of age in the past two centuries, this has not been matched by an improvement in healthspan [1]. In fact, age is one of the major risk factors for debilitating conditions such as cardiovascular and neurodegenerative diseases, having a large societal and economical impact [2, 3]. However, not all individuals age in the same way. In particular, sex seems to be responsible for a substantial inter-individual variability during aging, with women displaying a higher probability of developing certain age-related disorders such as Alzheimer’s disease [4] and multiple sclerosis [5], whereas men are more likely to develop Parkinson’s disease [6]. These differences in vulnerability to distinct diseases suggest that men and women have a distinct underlying brain network organization that might predispose them to develop specific pathological processes.

The functional network organization of the brain can be assessed using the coherence of spontaneous fluctuations in brain activity across brain regions by measuring the bloodoxygen-level-dependent signals on resting-state functional magnetic resonance imaging (rsfMRI) [7]. Using this technique, several studies have identified highly reproducible restingstate networks in the brain such as the sensorimotor, dorsal attention and default-mode networks, which play an important role in motor and cognitive functions [8]. The communication between these networks is particularly important for brain function and has been shown to change during the course of aging [9, 10], with older individuals showing a loss of anti-correlations (negative connections) [11, 12] and increases in positive correlations (positive connections) [13, 14] between resting-state networks. These changes reflect the tendency of older individuals to over-recruit functional networks, needing to activate more brain networks than younger individuals, thus decreasing functional specialization and spending more neural resources [15].

However, the impact of sex on the communication between functional brain networks during aging is still not well understood [16, 17], in part due to the small number of participants included in previous studies and their limited statistical power [17, 18]. Assessing sex differences in functional connectivity is important for several reasons. For example, because functional networks are closely associated with cognitive and sensorimotor functions [8], understanding how their communication deteriorates with aging might provide important clues on why men and women are vulnerable to different diseases [4–6] and why they show differences in other important health aspects such as brain structure, cardiovascular risk factors and cognitive function [17, 19–22].

From a methodological point of view, studies analyzing functional connectivity have mainly focused on positive connections [23, 24]. While this approach is more straightforward to assess the organization or topology of brain networks [25], negative connections are commonly found between brain networks or areas and seem to play an important role in brain communication [26, 27] and cognition [28]. Since the negative connections or anti-correlations carry behaviorally relevant information [28], an integrative approach that incorporates information from negative correlations as well as positive correlations may reveal unique insights on sex differences throughout aging.

In this study, we developed this approach by combining the positive and negative functional connections between networks as separate layers in a complex multilayer network. We demonstrated that the multilayer measures outperform traditional connectivity measures in predicting the different functional connectivity trajectories in men and women between 47 and 79 years old in a large cross-sectional cohort of 37543 individuals. Moreover, together with other measures of network organization, they were associated with structural brain imaging markers, cardiovascular risk factors, and cognitive functions, which typically differ between men and women [17, 19–22] as well as genes involved in physiological processes associated with aging. These findings open new avenues for the study of functional brain connectivity in aging using multilayer network measures.

## II. RESULTS

### Sample

We included 19975 women and 17568 men with resting-state functional MRI from the UK Brain Biobank cohort [29] (see *Methods*, section *Participants* and Supplementary Fig. S1). A subset of these individuals had available structural brain imaging data such as T1-weighted imaging and underwent a comprehensive assessment of cognitive functions and cardiovascular risk factors (see *Methods*, sections *Structural imaging preprocessing, Cardiovascular risk factors* and *Cognitive tests*. Using permutation testing to compare the demographic characteristics between women and men, we found that men showed higher scores than women in the executive cognitive domain during middle ages and in the visuospatial cognitive domain across all ages (Supplementary Fig. S2c-d). In line with previous research showing that men are more susceptible to cardiovascular problems during middle adulthood [30, 31], we observed a significantly greater prevalence of high blood pressure, heart attack and white matter hyperintensities in men between the ages of 51 and 76 compared to women (Supplementary Fig. S2e,f,k). Finally, men had larger subcortical volumes across all ages (Supplementary Figure S2i-k), consistent with previous studies [17, 32]. There were no significant differences between sexes in professional qualifications and years of education.

### Brain connectivity analysis

Functional brain connectivity was assessed for each participant using the negative and positive correlations between 21 nodes that correspond to the resting-state functional MRI networks shown in Fig. 1a and Supplementary Fig. S3 (see *Methods*, section *Functional image preprocessing*). First, we computed classical single-layer connectivity measures, namely, the average connectivity for the whole correlation network, followed by the average negative connectivity, the average positive connectivity and the number of negative correlations (see *Methods*, section *Connectivity measures*). Then, the negative and positive correlations of each functional network were separated (Figs. 1b.1 and 1b.2) and analyzed as two independent layers. To evaluate the topological organization of each single layer, we used two measures: the clustering coefficient and the global efficiency. The clustering coefficient is a measure of segregation that increases with the number of local connections and represents the average clustered connectivity around all nodes in the network. The global efficiency is a measure of integration that increases with the number of direct paths and estimates the average capability with which different nodes communicate with each other [33] (see *Methods*, section *Single-layer network measures*). Then, we integrated these two layers into a multiplex (Fig. 1c) and a multilayer (Fig. 1d) network to assess how they interact. In the multiplex network approach (Fig. 1c), each node in the positive layer was connected with the same node in the negative layer. We computed two multiplex measures: the multiplex clustering coefficient, a measure that increases with the local connections in neighboring nodes between the two layers, and the multiplex participation, which is a measure of integration that assesses how evenly a node is connected in the two layers [34] (see *Methods*, section *Multiplex network measures*). A disadvantage of the multiplex approach is that the interaction between the two layers is local being only allowed between the same nodes. To address this limitation, in the multilayer network approach we connected each node in one layer to every node in the other layer (Fig. 1d). The interaction strength between the two layers can be changed by adjusting the weight of the interlayer connections, *σ*. For each multilayer network, we define *σ* as a fraction of the strongest functional connections in the corresponding network, and evaluate the measures’ ability to characterize sex differences across the wide range of *σ* values. We developed two new measures to assess the integration and segregation properties of these multilayer networks. Specifically, we calculated the multilayer global efficiency, which compares the global efficiency differences due to the intra- and interlayer connections. Similarly, we also calculated the multilayer clustering coefficient, which compares the clustering coefficients or triangles in all nodes between the two layers (see *Methods*, section *Multilayer network measures*).

**FIG. 1.**
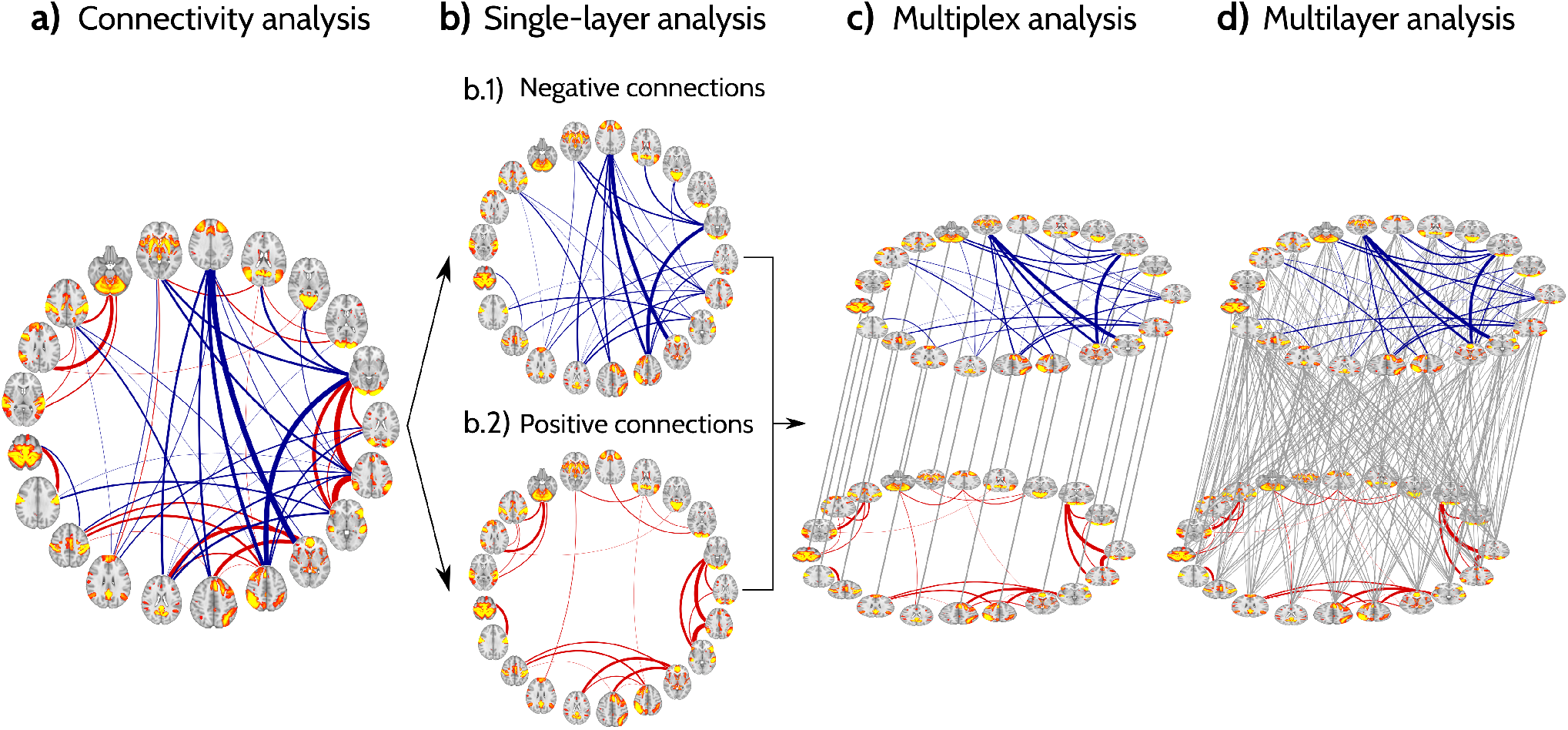
Analysis workflow. a) Example of the 21 resting-state networks used as nodes and their positive (red) and negative connections (blue) for one of the subjects included in the analyses. b) The positive and negative connections were split into two networks: one negative (b.1) and the other one positive (b.2). The topology of these two networks was evaluated using the clustering coefficient and global efficiency. c-d) The positive and negative networks were then integrated as two separate layers in a multiplex network (c) (where each node in one layer is connected to the same node in the other layer) and a multilayer network (d) (where each node in one layer is connected to all other nodes in the other layer). We evaluated the topology of the multiplex network using the clustering and participation coefficients, whereas the topology of the multilayer network was assessed using the novel multilayer global efficiency and multilayer clustering coefficient. In all graphs, thicker connections represent stronger positive or negative functional connections.

### Women have less negative connections than men

In a first step, to identify which simple connectivity measures showed the greatest differences between sexes over the course of aging, we compared men and women at all ages using permutation test and used separate linear models that included whole-brain average connectivity, average negative connectivity, average positive connectivity and the number of negative correlations as the outcome and age, sex, age^2^, age×sex and age^2^×sex interactions as predictors (see *Methods*, section *Statistical analysis*). These models showed that women had significantly higher average functional connectivity than men (R^2^ = 0.769; AIC = 244.651; MSE = 2.184; Fig. 2a and Supplementary Fig. 4a), whereas men had a significantly higher number of negative connections compared to women (R^2^ = 0.778; AIC = 501.106; MSE = 108.420; Fig. 2d and Supplementary Fig. 4d) across a broad age range (50 to 71 years). These differences in connectivity between sexes diminished with increasing age, to the point where there were almost no differences in mean connectivity strength or number of negative connections between men and women after 75 years (age×sex interaction: average connectivity p = 0.01; number of negative connections p *<* 0.001). On the other hand, there were no significant differences between sexes in the average positive and negative connectivity strength (R^2^ = 0.403; AIC = 281.229; MSE = 3.947 and R^2^ = 0.408; AIC = 281.229; MSE = 2.013 respectively; Fig. 2b-c and Supplementary Fig.4b-c). These results suggest that women have higher connectivity strengths than men due to a lower number of negative connections, but these differences dissipate with increasing age.

**FIG. 2.**
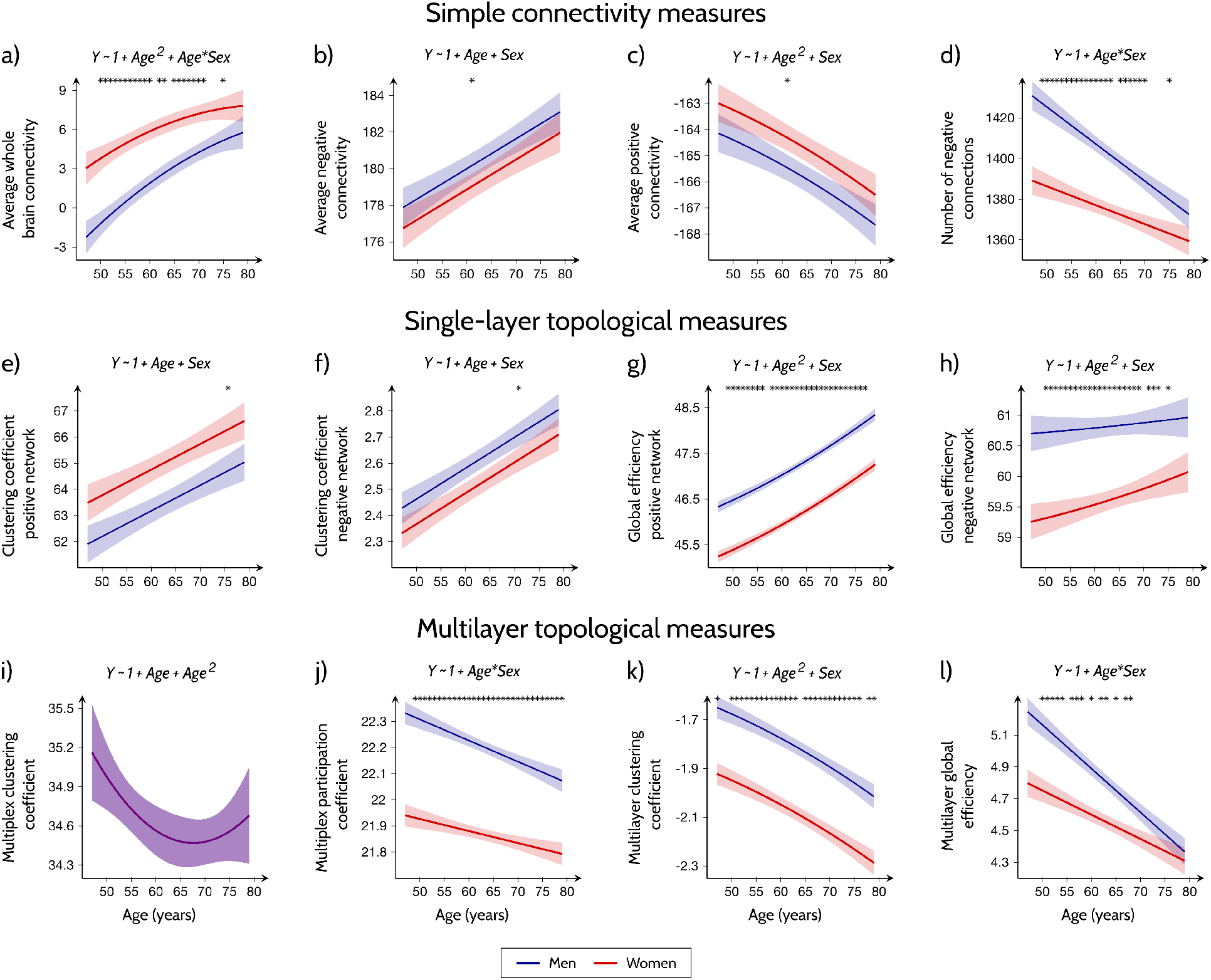
Prediction of functional connectivity. Results of the linear models with simple connectivity measures (a-d), single-layer topological measures (e-h) and multi-layer topological measures (i-j, multiplex; k-l, multilayer) as the outcomes and age, sex, age^2^, age×sex and age^2^×sex interactions as predictors. The areas show the 95% confidence intervals (CI) for the predictions and the solid lines show the best line fit. The stars indicate points that showed significant differences between men and women after correction for multiple comparisons across the different age groups (FDR at q *<* 0.05). Details about the models are summarized in Supplementary Table S1 and the Statistical analysis section.

### Men have shorter network paths than women

To identify which single-layer topological measures showed the greatest differences between sexes over the course of aging, we repeated the above analysis by including them as dependent variables in separate linear models with age, sex, age^2^, age×sex and age^2^×sex interactions as predictors. These models showed that men had higher global efficiency than women in the positive and negative layers (R^2^ = 0.924; AIC = −1.051; MSE = 0.055 and R^2^ = 0.649; AIC = 88.603; MSE = 0.209 respectively; Fig. 2g-h and Supplementary Fig. 4g-h), indicating that their functional connectomes were characterized by shorter paths in the networks with negative and positive connections. Interestingly, these sex differences remained constant across different ages, suggesting that they were independent of age (age×sex or age^2^×sex interaction not significant in global efficiency measures, Supplementary Table S1). In contrast, no significant differences in the clustering coefficients in the positive and negative networks were observed between women and men (R^2^ = 0.459; AIC = 226.729; MSE = 1.728 and R^2^ = 0.533; AIC = −96.294; MSE = 0.013 respectively; Fig. 2e-f and Supplementary Fig. 4e-f).

### Men have a greater balance between positive and negative connections

Our linear regression models showed that the multiplex participation coefficient was significantly higher in men than in women across all ages (R^2^ = 0.891; AIC = −172.848; MSE = 0.004; Fig. 2j and Supplementary Fig. 4j), indicating that the men’s functional connectomes were characterized by nodes with a similar number of connections in the negative and positive connectivity layers. However, these differences decreased with age, similarly to the average connectivity and number of negative connections (age×sex interaction: p = 0.036). The multiplex clustering coefficient did not reveal any significant differences between men and women (R^2^ = 0.096; AIC = −106.989; MSE = 0.282; Fig. 2i and Supplementary Fig. 4i).

### Multilayer topological measures are lower in women than in men

The differences between men and women were robust for the complete range of interlayer weights *σ* for the multilayer clustering and for smaller values of *σ* in the case of multilayer global efficiency. However, the strongest differences were observed for the *σ* = 0.7 and *σ* = 0.2 in the case of multilayer clustering and multilayer global efficiency respectively (Supplementary Figs. S5 and S6). Whereas the multilayer clustering coefficient differences between men and women remained stable with aging (R^2^ = 0.799; AIC = −130.497; MSE = 0.008; age×sex interaction not significant; Fig. 2k and Supplementary Table S1), the differences in multilayer global efficiency decreased with aging (R^2^ = 0.795; AIC = −82.061; MSE = 0.016; age×sex interaction: p *<* 0.001; Fig. 2l and Supplementary Table S1). Up to 80% of the variance in both multilayer measures was associated with sex differences over time, outperforming all previous models for simple connectivity, single-layer and multiplex topological measures. These findings suggest that complex network measures that account for a full interaction between positive and negative functional connections are more sensitive to sex differences across aging.

### Multilayer and multiplex measures are the best predictors of structural, cognitive and cardiovascular differences between men and women

Next, we assessed whether the observed sex differences in functional connectivity throughout aging were associated with the cognitive functions, structural brain measures and vascular risk factors that differed between men and women in our cohort (executive functions, visuospatial functions, blood pressure and heart attack prevalence, subcortical volumes, white matter hyperintensities; see *Results*, section *Sample* and Supplementary Fig. S2). Due to the high collinearity between the functional connectivity measures, we examined these associations using partial least squares (PLS) regressions. The executive cognitive scores were best explained by the single-layer positive global efficiency, followed by the single-layer negative clustering, average positive connectivity and negative connectivity. The visuospatial cognitive scores were best explained by the multilayer functional connectivity measures, followed by the average connectivity, number of negative connections and the positive single-layer global efficiency. Heart attack and high blood pressure prevalence were best predicted by multiplex participation and single-layer global efficiency of the positive connections, with blood pressure additionally being predicted by multilayer clustering coefficient, number of negative connections, average connectivity and single-layer global efficiency of the positive network. Multilayer global efficiency was a significant predictor of subcortical brain volumes and white matter hyperintensities, in addition to the average connectivity, number of negative connections, multilayer clustering coefficient, multiplex participation and single-layer clustering coefficient in the case of subcortical volumes and single layer clustering coefficient in the case of white matter hyperintensities. The detailed VIP scores for each predictor and the PLS model cumulative explained variance for the predicted variables are shown in Fig. 3 and Supplementary Tables S2-S7. Altogether, these results indicate that, compared to the other measures, the multilayer and multiplex measures were the best predictors of cognitive functions, structural brain measures and vascular risk factors that differed between men and women during aging.

**FIG. 3.**
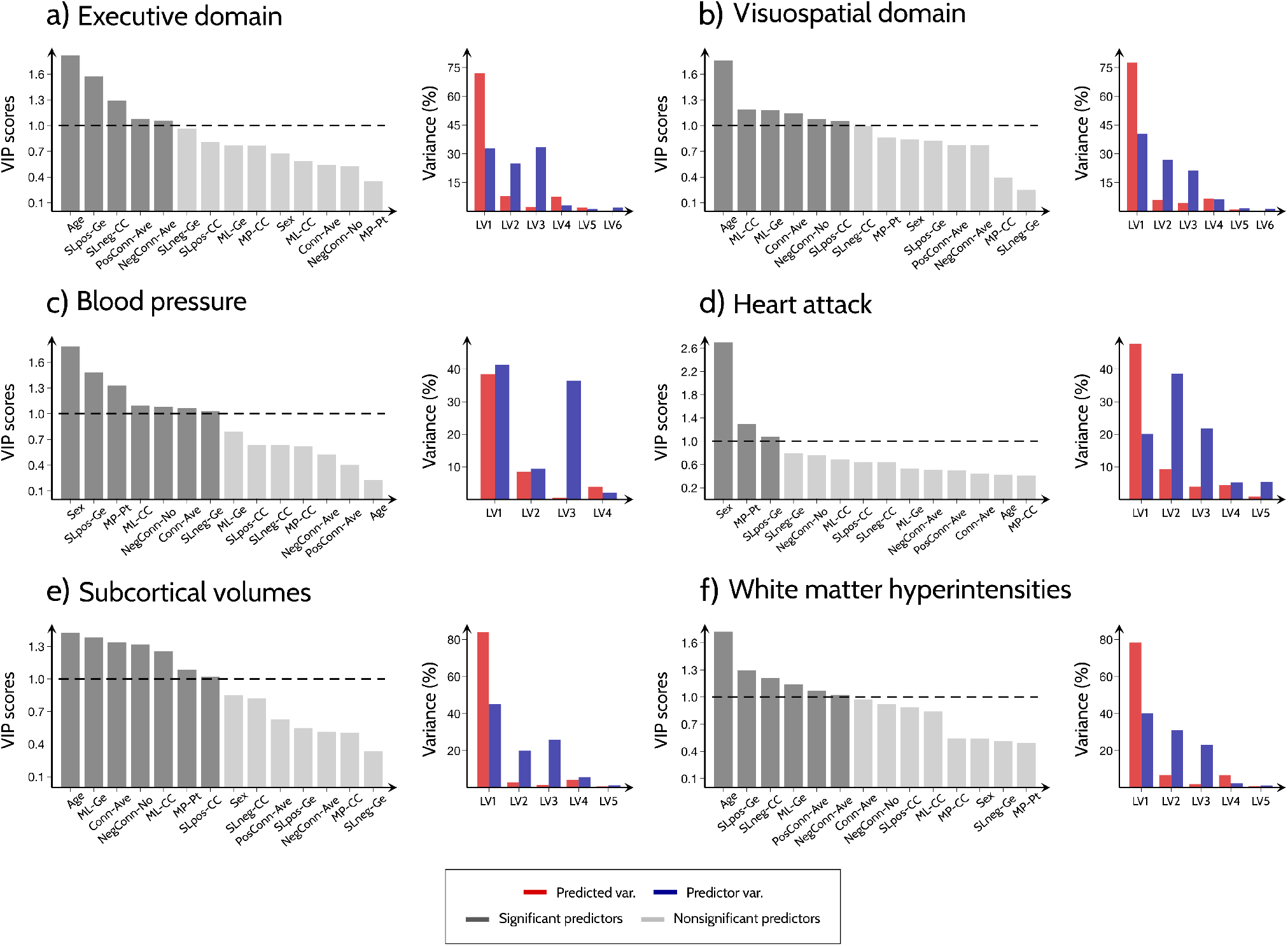
Relationship between functional connectivity measures and brain structure, cognitive measures and cardiovascular disease factors. Plots showing the VIP scores for all functional connectivity measures and the amount of variance (for predicted and predictor variables) explained by the corresponding latent variables. The PLS analysis was performed for measures showing significant differences between men and women in: a) executive and b) visuospatial domains; c) prevalence of high blood pressure and d) heart attack; e) subcortical volumes and f) white matter hyperintensities. Abbreviations: LV: latent variables; Conn-Ave: Average connectivity; PosConn-Ave: Average positive connectivity; NegConn-Ave: Average negative connectivity; NegConn-No: Number of negative connections; SLpos-CC and SLneg-CC: Single layer clustering coefficient for networks of positive and negative connections; SLpos-Ge and SLneg-Ge: Single layer global efficiency for networks of positive and negative connections; MP-CC: Multiplex clustering coefficient; MP-Pt: Multiplex participation coefficient; ML-CC and ML-Ge: Multilayer clustering coefficient and global efficiency.

### Multilayer and multiplex measures are associated with genes involved in agingrelated physiological processes

To identify genetic variation associated with the 12 multilayer, single-layer and connectivity functional measures we examined 9,356,431 genetic variants with minor allele frequency > 1% in a total of 33 773 European participants (see *Methods*, section *Genetic association analyses*). A total of 4 *loci* exceeded a genome wide significance level among one or more of these traits (Fig. 4 and Supplementary Figure S7). A locus near Paired Box 8 (*PAX8*), a gene involved in sleep efficiency, diastolic blood pressure, development and vulnerability to neurodegenerative diseases [35–37], showed an association with the majority of functional connectivity measures, including multilayer and multiplex clustering, as well as all single-layer measures. Colocalization analysis with eQTL summary statistics of 49 tissues in GTEx project and brain tissues in BrainSeq, ROSMAP, Braineac2 and CommonMind datasets (see *Methods*, section *Colocalization*) [38–43], suggests a consistent strong colocalization between this locus and gene expression patterns of Immunoglobulin Kappa Variable 1/OR2-108 (*IGKV1OR2-108*), COBW domain-containing protein 2 (*CBWD2*) and Forkhead Box D4 Like 1 (*FOXD4L1*) over the multilayer and multiplex clustering coefficients, average positive and negative connectivity and all single-layer functional measures (Supplementary Table S8). Furthermore, a locus near Inositol Polyphosphate-5-Phosphatase A (*INPP5A*) gene showed significant associations with the single-layer positive global efficiency. Interestingly, the genome-wide association study (GWAS) lead variant at this locus colocalized with eQTL for *INPP5A* in brain dorsolateral prefrontal cortex (DLPFC) from CommonMind dataset. Finally, loci near Adenosine Deaminase RNA specific B2 (*ADARB2*) and D-Amino Acid Oxidase Activator (*DAOA*) genes, which have previously been linked with processes involved in normal memory functioning and ATP metabolism [44] were associated with multiplex clustering and multiplex participation coefficients respectively.

**FIG. 4.**
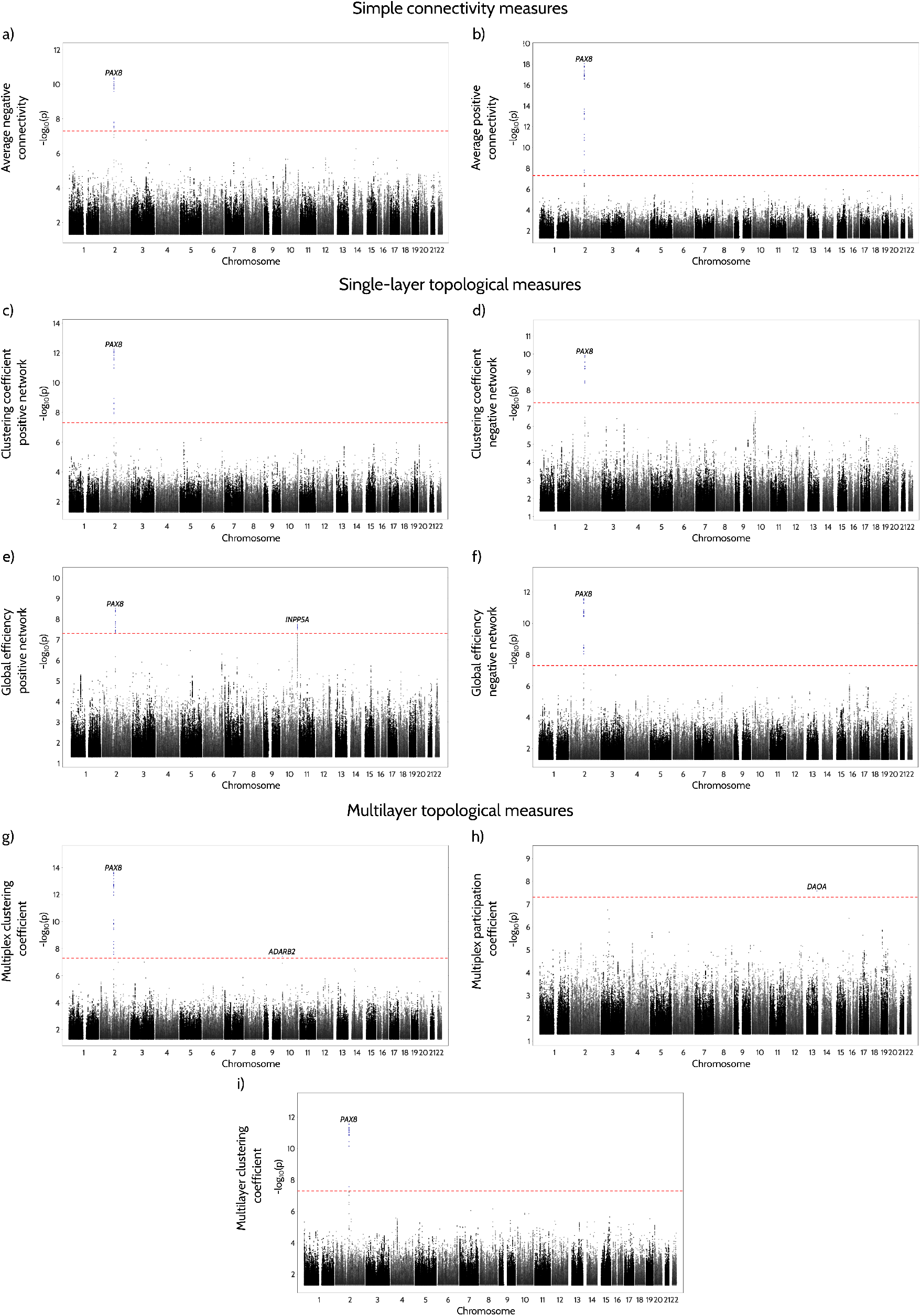
Manhattan plots for GWAS analysis of functional connectivity measures. The results from the GWAS analysis of simple connectivity measures (a-b), single-layer topological measures (c-f) and multi-layer topological measures (g-h, multiplex; i, multilayer) in a total sample of 33 773 individuals. The y axis shows the p-values for the association tests on a -log10 scale, while the different chromosomes (differentiated by the black and gray colors) are ordered on the x axis. The lead variants that surpass the genome-wide significance threshold (indicated by the red line) are highlighted as blue circles

## III. DISCUSSION

Complex network measures are becoming increasingly popular in the field of network science [8]. In combination with large samples, these measures can improve our understanding of brain connectivity [25], and, in particular, our ability to predict sex differences over the course of aging [45]. Previous studies of brain connectivity and network topology have mainly focused on measures that exclude the negative connections or average the effects of positive and negative connections [13, 23, 46–48]. However, changes in the interaction between positive and negative connectivity could be a more sensitive marker of abnormalities that occur in men and women in middle and late adulthood. Here we show that measures that assess this interaction are indeed better predictors of sex differences during aging. Furthermore, we also show that these measures are associated with genes implicated in aging-related physiological processes as well as cognition, brain structure and vascular disease, which have been previously shown to differ between men and women [17, 19–22]. Altogether these findings highlight the importance of integrating the information from positive and negative connections in a multiplex or multilayer network approach to provide a more holistic view of functional brain connectivity changes. Advancing age is associated with lower physical fitness and worse cognitive abilities, being one of the greatest risk factors for the development of neurodegenerative diseases [49, 50]. However, there are several factors that determine how an individual ages and his/her predisposition to develop certain diseases. Sex is one of these factors and it has recently received a lot of attention due to increasing recognition that precision medicine approaches should take into account biological sex to treat neurodegenerative diseases [51]. Although many studies have described how men and women age differently [52], the location and nature of these differences in functional brain connectivity vary across studies [16]. Functional brain connectivity plays a crucial role in how brain networks communicate with each other, being closely associated with behavior and cognition [8]. Previous studies assessing connectivity between resting-state networks have shown an increase in the connections between these networks with aging [10, 13, 14, 23]. These increases in connectivity are thought to be due to a less efficient use of neural resources in older individuals, who tend to over-recruit brain networks to compensate for the detrimental effects of aging [15, 53, 54]. Here, we confirm these findings in a larger sample of middle-age and old adults by showing that aging is associated with increases in the average whole brain connectivity and decreases in the number of negative connections. Moreover, our findings revealed that men displayed lower average connectivity and a greater number of negative connections than women at young ages but these differences dissipated with increasing age, to the point that no differences between sexes were observed any longer at old ages. These findings agree with results from previous studies showing different functional trajectories between men and women [16, 55]. However, in contrast to previous studies, here we demonstrate that, although the functional connectomes of younger men and women are different, they become increasingly similar with older age possibly due to a faster rate of functional changes observed in the brains of men [48], which is in line with studies showing that men have lower resilience to age-related cognitive decline compared to women [22].

Regarding measures of network organization, we found that the positive global efficiency was higher in men compared to women but these differences remained stable across different ages. Similar differences were found in the negative global efficiency; however, in this case, the differences were not stable and seemed to decrease with aging. The global efficiency is used to assess integration or the ability for an efficient processing to occur between distant brain networks [25]; an excessive integration is thought to impede the ability of the brain to process information in a meaningful way [56]. Furthermore, global efficiency is associated with the presence of long range connections between networks. The healthy brain connectome is characterized by a low number of such connections because long range connections are associated with higher metabolic costs and minimizing these costs is essential for evolution [57]. Therefore, these differences between sexes might provide clues on why men are more prone to develop specific diseases with age, for example Parkinson’s disease and epileptic seizures, which are associated with abnormal organization in the functional connectivity networks as a result of increased integration [56, 58].

To evaluate whether the interaction between the positive and negative connections can reveal additional insights into sex differences over the course of aging, we integrated these connections as two separate layers in a multiplex and a multilayer networks. In the multiplex network, where only connections between the same nodes in the two layers are allowed, we found that men had higher multiplex participation coefficients compared to women. This result indicates that men had a higher balance between positive and negative connections in the two layers, which decreased with aging at a faster rate compared to women. Because the positive and negative connections can arise from different neurovascular mechanisms [59], the presence of both connection types is necessary to reach a more balanced communication between resting-state networks and consequently a better cognitive performance [60]; in fact, the lack of anti-correlation or negative connections has been associated with lower cognitive control and working memory performance during neurodevelopment [61]. Consistent with these findings, our results suggest that the less efficient connectomes reported in older persons may be related to an increase in the number of positive connections at the expense of negative connections [13], thereby disrupting this balance.

In the multilayer network, where connections are allowed between all nodes in the two layers, we found that men had higher multilayer clustering and multilayer global efficiency compared to women, which both decreased with aging. The multilayer clustering reflects the local connections that form triangles between the nodes of the two layers, whereas the multilayer global efficiency reflects the direct paths between the nodes in the two layers. These findings indicate that the positive and negative layers interact with each other to a higher degree in men than in women. This higher interaction might come at the expense of greater neural resources and metabolic costs, which could predispose the male connectome to the effects of increased oxidative stress and poor antioxidant defense mechanisms, which have been suggested to accompany higher brain connectivity [62] and could potentially lead to steeper rates of cognitive decline [22].

When the different connectivity and topological network measures were compared to each other, we found that the multiplex and multilayer network measures were the variables that were best predicted by sex differences over aging. In particular, age and sex were able to predict multiplex participation coefficient, multilayer clustering and multilayer global efficiency by explaining up to 89.1%, 79.9% and 79.5% of the variance. These findings indicate that the integration of positive and negative connections as separate layers in a complex network approach is sensitive to important age and sex-related variability not captured by conventional measures. This approach could thus be used to understand why men and women age differently. For instance, we found that cardiovascular risk factors such as hypertension and heart attack prevalence, which were higher in men between 51 and 79 ages, were best predicted by these measures. In addition, differences between sexes that remained stable over aging such as lower visuospatial cognitive scores and lower subcortical volumes in women compared to men were also best explained by multilayer measures.

Regarding the results of the GWAS analysis, a locus near *PAX8* gene showed an associations with the majority of functional connectivity measures. Interestingly, this locus has been associated with sleep efficiency, diastolic blood pressure, insomnia, and sleep length [37]. These data are consistent with the aging process resulting in changes in sleep habits and high blood pressure [20, 63], both of which have been linked to functional connectivity changes [35, 64, 65]. The *PAX8* protein has also been associated to the regulation of multiple genes involved in thyroid hormone synthesis, which is necessary for brain development and function, for example, through processes such as neuronal differentiation, synaptogenesis, and dendritic proliferation [35]. All functional measures associated with the *PAX8* gene demonstrated substantial colocalization with the *FoxD4L1* gene, which is similarly implicated in processes that promote the onset of neural differentiation [66]. They also demonstrated colocalization with *CBWD2*, which has been linked to sleep duration [35], and *IGKV1OR2-108*, which has been found to be elevated in the livers of type 2 diabetes patients [67] and can lead to abnormal functional connectivity [68].

The *rs4309079* locus, associated with single-layer positive global efficiency, has been linked to functional connectivity measures in previous research [36]. It is located adjacent to the *INPP5A* gene, which is involved in calcium signaling. As a fundamental cellular mechanism, inositol calcium signaling is expected to play a role in a range of neurological pathways that underlie functional connectivity [64]. The *INPP5A* has been further associated with brain age explained by changes in functional connectivity and decreased system segregation [64, 69], which is consistent with our current findings.

Multiplex participation showed a strong association in a locus near *DAOA* gene, which is known to play an important role in the control of glutamatergic transmission [44]. Glutamate is the most common excitatory neurotransmitter in mammals, and glutamate’s activation of NMDA receptors is critical for normal memory function. Glutamatergic antagonists (e.g., ketamine) have been demonstrated to lower performance on tests of declarative memory, verbal fluency, and problem solving, all of which have been linked to aging [44]. We nevertheless, could not find any striking colocalization evidence in the examined gene expression QTL datasets. Finally, for multiplex clustering, we found a locus near *ADARB2* (index variant: *rs2152237*) gene, which is involved in ATP/ITP metabolic pathway. As a result, our findings add to our understanding of the genetic influences on functional connectivity and provide a link between the functional and genetic architecture of the brain, which might be relevant in explaining the changes in a variety of biological processes throughout healthy aging.

This study has some limitations. First, a longitudinal study design would have been more appropriate to assess age-related differences between sexes in functional brain connectivity. However, although many efforts to collect large longitudinal samples are currently underway [29], unfortunately this data is not yet available. Thus, at the moment, we can only infer longitudinal changes based on large cross-sectional data sets and our findings must be interpreted with this limitation in mind. Second, there was a considerable overlap between the values for individual men and women in functional connectivity measures. This overlap has also been observed in previous studies [17, 45, 47, 70, 71], indicating that the prediction of an individual’s sex from a small number of functional connectivity measures is challenging [71]. Nonetheless, our findings revealed consistent group differences between men and women across a wide age range (47 - 79 ages), suggesting that, although there is considerable variability, some changes seem to be quite robust [72]. Finally, since in this study the nodes of the networks corresponded to 21 resting-state networks derived from independent component analyses [29], the negative connections are related to anti-correlations between resting-state networks, which have been consistently observed and demonstrated to have a neurophysiological basis [27, 73, 74]. Thus, these connections should be interpreted differently than the negative correlations between brain regions analyzed in studies using brain areas as nodes, which are still not clearly interpretable [25, 74].

To summarize, in this study we developed novel multilayer connectivity measures in order to assess the connectivity patterns and topological architecture between resting-state networks in a large cohort of middle and old adults. We showed that these multilayer measures are superior at capturing sex-related effects during aging when compared to simpler connectivity measures that do not account for the interaction between positive and negative connections. The multilayer measures were also significant predictors of sex differences in cognitive, structural and cardiovascular measures and they were associated with genes that have previously been implicated in aging-related processes. Thus, our findings highlight the importance of studying the interaction between positive and negative functional connections to understand the effects of sex over aging, which should be included in future studies.

## IV. METHODS

### Participants

The UK Biobank cohort is a large population-based study with more than 500,000 participants from the United Kingdom (https://www.ukbiobank.ac.uk/). Following an initial visit for collection of medical and other clinical information, 37704 individuals underwent MRI. To achieve robust group comparisons, we limited our analyses to age groups with at least 50 participants, resulting in a sample size of 37543 people in the age range 47-79 years (17568 men: mean age = 64.75; std = 7.58 and 19975 women: mean age = 63.49; std = 7.33).

### Image acquisition

The functional MRI scans were performed on a standard Siemens Skyra 3T scanner using an echo-planar imaging (EPI) sequence with the following parameters: duration ∼ 6 min; 490 timepoints; repetition time = 735 ms; echo time = 39 ms; field of view = 88 × 88 × 64; voxel size = 2.4 mm^3^; flip angle = 52^º^. Participants were instructed to relax and think of nothing in particular while focusing their eyes on a crosshair during the scan. Regarding structural MRI, the T1-weighted images were obtained using a 3D magnetization-prepared rapid gradient-echo imaging sequence with the following parameters: 208 slices, echo time = 880 ms; repetition time = 2000 ms; field of view = 208 × 256 × 256; voxel size = 1 mm^3^ [75].

### Functional image preprocessing

The functional MRI scans were analysed using an image-processing pipeline run by the UK Biobank imaging core. After data acquisition, a number of pre-processing steps were carried out using FSL, including motion correction using MCFLIRT, grand-mean intensity normalization by a single multiplicative factor, high-pass temporal filtering with a Gaussian-weighted least-squares straight line fitting (*σ* = 50.0s), EPI unwarping by using a field map obtained before data collection, gradient distortion correction (GDC) unwarping, and removal of all artifacts by an ICA-based X-noiseifier. Finally, all datasets underwent temporal demeaning and variance normalization.

The pre-processed data of 4100 participants was used for a Group-ICA analysis. Using FSL’s MELODIC tool and FSLNets toolbox (https://fsl.fmrib.ox.ac.uk/fsl/fslwiki), a spatial-ICA with a dimensionality of 25 components was applied and the resulting ICA maps were mapped onto each subject’s resting-state fMRI timeseries data to generate one representative timeseries per ICA component. During this procedure, four networks were identified as artifacts and were discarded from further analysis. This resulted in a 21 × 21 connectivity matrix for each participant, where the functional connectivity between each pair of ICA spatial maps is characterized by full normalized temporal correlation. Analysed networks included the default-mode, frontoparietal, salience, sensorimotor, visual, auditory, subcortical and cerebellar networks; the group-ICA spatial maps can be found at: https://biobank.ctsu.ox.ac.uk/crystal/refer.cgi?id=9028 and the average functional connectomes for several representative age groups are shown in Supplementary Fig. 2. The complete description of the pre-processing procedures can be found elsewhere [29, 75] (Online Documentation: http://biobank.ctsu.ox.ac.uk/crystal/docs/brainmri.pdf).

### Structural imaging preprocessing

T1-weighted scans were preprocessed using the standard procedures of the FreeSurfer pipeline (version 6.0; https://surfer.nmr.mgh.harvard.edu/). We calculated the cortical thickness for each individual by averaging the regional cortical thicknesses from 68 regions from the Desikan-Killiany atlas [76]. In addition, we calculated the subcortical volumes for each individual by averaging the volumes of all subcortical gray matter structures (cerebellar cortex, thalamus, caudate, putamen, pallidum, hippocampus, amygdala, accumbens), which were corrected for total intracranial volume using a regression approach [77]. This data was available for a subsample of 17317 men and 19842 women.

### Cardiovascular risk factors

At their initial visit, individuals were questioned about their medical history, including whether or not they had a high blood pressure (subsample of 17517 men and 19911 women), a heart attack (subsample of 15204 men and 16918 women), a heart attack-related angina episode (subsample of 14224 men and 15520 women), or a stroke (subsample of 14224 men and 15520 women). We calculated the percentage of men and women diagnosed with the previous conditions at all age groups, and used these percentages as dependent variables in further analyses. Since the diagnosis of cardiovascular diseases was performed at a different timepoint than the brain scanning, we included the time difference between diagnosis and scanning as a covariate in the analyses with cardiovascular variables. Furthermore, as there were several age groups that did not have any individuals with heart attack and hypertension, the models for these variables were corrected for age prior to the PLS regression.

### Cognitive tests

The cognitive assessments were administered on a touch screen and took place at the same visit as the brain scans. We averaged the standardized z-scores of 10 cognitive tests [78] into 4 different cognitive domains: attention/psychomotor speed (reaction time, trail making - numeric path, symbol digit substitution tests; subsample of 11531 men and 13125 women), memory (numeric memory, paired associative learning, prospective memory, pairs matching tests; subsample of 11657 men and 13284 women), executive (fluid intelligence/reasoning, trail making - alphanumeric path tests; subsample of 11535 men and 13159 women), and visuospatial/visuoconstructional (matrix pattern completion, tower rearranging tests; subsample of 11452 men and 13013 women). More details about the cognitive tests are available at: https://biobank.ndph.ox.ac.uk/showcase/label.cgi?id=100026.

### Connectivity measures

We assessed the functional connectomes using 4 different connectivity measures. A network’s average connectivity (Conn-Ave) is defined as the average functional strength of all its connections. Similarly, we estimated the average positive connectivity (PosConn-Ave) and average negative connectivity (NegConn-Ave) as the mean strength of the network’s positive and negative connections, respectively. Finally, we calculated the number of negative connections NegConn-No in the network. These measures can be evaluated as:

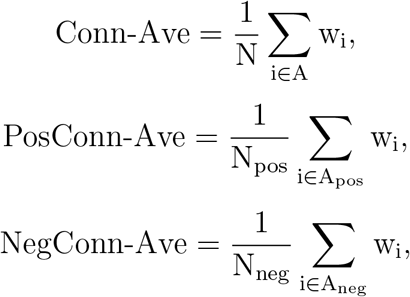

where N is the total number of connections in the network A. The network A can be expressed as a sum of A_pos_ and A_neg_, which denote the networks consisting of only positive and negative connections. The total number of connections in A_pos_ and A_neg_ are denoted by N_pos_ and N_neg_ respectively.

The connectivity measures were calculated on the weighted connectivity networks consisting of positive and negative connections (shown in Fig. 1a). For each weighted connectivity network, we calculated a corresponding binary network in which the individual connections retained their weight if they exceeded a certain threshold and were set to zero if they did not. In this process, the absolute value of each connection was compared to the threshold, however, their sign was preserved in the resulting binary matrix (i.e., negative connections in the weighted network remained negative in the binary network). As there are multiple thresholding approaches and there is currently no consensus as to which network density should be used [25], we performed the thresholding at a density range of 6% to 33%, in steps of 1%. For densities below 6%, the networks became largely disconnected with fewer edges than nodes, whereas 33% was the maximum density that could be reached by all men’s and women’s networks of positive and negative connections (Supplementary Figure S8).

After calculating all measures at each density within the complete density range, we evaluated the corresponding area under the curve (AUC) value, which was used to assess the between-sex differences (Supplementary Fig. S9). The AUC value was obtained by numerically integrating the measure values over the density range; this procedure resulted in a single numerical value for each network measure across the range of densities. As this analysis takes into account the entire density range, it is considered to be less susceptible to the thresholding process [25].

### Topological network analysis

#### Single-layer network measures

We split the weighted connectivity networks into networks consisting of positive connections and negative connections (Fig. 1b). As network measures are not defined for negative weights, the weights in the network of negative connections were substituted by their absolute values. Therefore, the topological analysis was equivalent for both positive and negative connections networks; in the following we omit the subscript (pos/neg) for clarity. Both networks were independently binarized at the density range 6% - 33%, and the AUC value for all measures was obtained across this range, as outlined above. The topology of these networks was evaluated by calculating the global efficiency (SL-Ge) and clustering coefficient (SL-CC).

#### Single-layer global efficiency

In a weighted network, the distance d_ij_ between nodes i and i is the total sum of individual connection lengths along the shortest path that connects the two nodes. Because large weights often imply strong relationships and close proximity, connection lengths are inversely proportional to connection weights. While the shortest paths have the smallest weighted distance, this does not necessarily equate to having the fewest number of edges [33]. The regional global efficiency of a node i, denoted by SL-Ge_i_, is defined as the average inverse distance from i to the other nodes in the network. The global efficiency of a network, SL-Ge, is determined as the average of the global efficiency of all nodes:

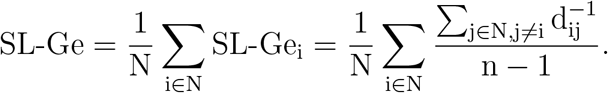

#### Single-layer clustering coefficient

The clustering coefficient of a given node represents the fraction of the total number of triangles that are present around it. The network clustering coefficient is derived by averaging the clustering coefficients of all nodes. Using the definitions of the number of triangles and nodal degree, the network clustering coefficient can be represented as [33]:

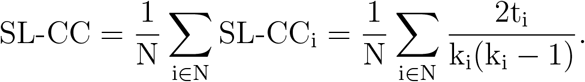

k_i_, the degree of a node i, is defined as the total number of connections i has with other nodes in the network, regardless of their weight. If the neighbors of node i are connected with each other, a triangle can be constructed around i. In weighted networks, the total number of triangles around node i, t_i_, is calculated by summing the contributions of each individual triangle, defined as the geometric mean of the triangle’s edge weights:

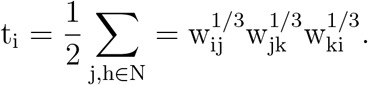

#### Multiplex network measures

For each individual, we built a multiplex network with two layers, one with positive connections and the other with negative connections (Fig. 1c). The multiplex networks were calculated at densities ranging from 6% to 33% by combining the two single-layer networks at the respective densities. At each density the multiplex networks can be represented by a supra-adjacency matrix, W, which consists of the intra-layer adjacency matrices on the main diagonal and the inter-layer connections in the off-diagonal entries. The inter-layer connections are formed only among the node’s replicas, i.e. 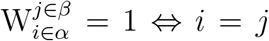, where *α* and *β* denote layer 1 and layer 2 respectively. We evaluated the topology of these networks by calculating the participation and clustering coefficients and evaluating the AUC over the complete density range.

#### Multiplex participation coefficient

This measure, MP-Pt, is used to quantify the heterogeneity in the connectivity patterns of a node i across the different layers. It is calculated as:

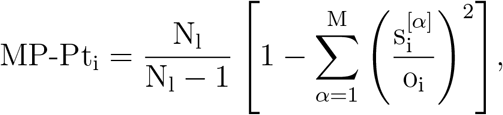

where N_l_ is the number of layers and 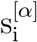 is the strength of node i at the *α*-th layer, defined as the sum of the weights of all edges connected to i. Finally, 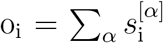 is the overlapping strength of the node i. MP-Pt_i_ determines whether i’s connections are evenly spread throughout the layers or are largely concentrated in one or a few layers. MP-Pt_i_ has values in the range 0 to 1 and, in general, bigger MP-Pt_i_ values suggest a more equal distribution of node i’s connections in all layers of the multiplex network. The global participation coefficient MP-Pt of the multiplex network is defined as the average of nodal MP-Pt_i_ coefficients over all nodes.

#### Multiplex clustering coefficient

The clustering coefficient can be used to assess whether the clustering features of the aggregated multiplex network differed from those of the individual layers. In contrast to single-layer clustering, a triangle in a multiplex network is formed by one edge from one layer and the remaining two edges from the other layer. Therefore, the multiplex clustering coefficient of a node i (MP-CCi) is defined as:

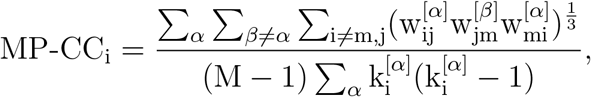

where 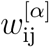 denote the connection between the nodes i and j in the layer *α* and k_i_ denotes the degree of a node i. The global multiplex clustering coefficient is calculated as the average of nodal clustering coefficients across all nodes.

#### Multilayer network measures

We constructed two-layer multilayer networks for each individual following an analogous procedure as the one of multiplex networks. However, in contrast to the multiplex approach, we established weighted inter-layer connections between all node pairs to quantify the degree of interaction between the two layers. In this framework, the level of between-layer interactions is an independent variable that can be controlled by the weight of the inter-layer connections. Due to the variability of the intra-layer connection weights in all subjects, we define the inter-layer weights, w_ij_, as a fraction of each participant’s biggest absolute functional connection:

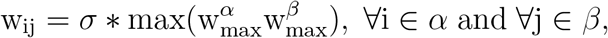

where 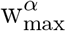 and 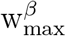 denote the maximum weight within layers 1 and 2 respectively, and *σ* is the fraction of the maximum weight. In our analyses, we have evaluated the multilayer in the *σ* range of 0.05 to 1, in steps of 0.05.

#### Multilayer global efficiency

We developed a new measure of multilayer global efficiency for each node i, denoted by ML-Ge_i_, which reflects the difference between the average inverse distances from i to all nodes in layer 2 and layer 1. Therefore, to evaluate ML-Ge_i_, we first computed the global efficiency of node i (as defined in section Single-layer global efficiency.) by considering only the shortest paths from i to nodes in the layer 1 and nodes in layer 2 separately, denoted by 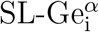 and 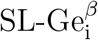. Then, ML-Ge_i_ can be defined as:

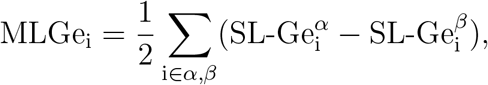

where the summations run over the two replicas of node i in the two layers. Therefore, ML-Ge_i_ measures the difference in global efficiency between the layer of negative and positive connections at different levels of inter-layer interaction that depends on the value of *σ*. This interaction is reflected through the values of 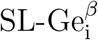; different inter-layer strength result in different global efficiency values between i and nodes in the layer not containing i. Smaller values of ML-Ge_i_ suggest that the two layers have more comparable integrative qualities, whilst higher values indicate that the two layers have a more divergent topology in terms of information integration. The network multilayer global efficiency can be defined as the average of ML-Ge_i_ calculated across all nodes in the network.

#### Multilayer clustering coefficient

We defined this measure analogously to ML-CC_i_, in order to quantify the difference in the clustering properties between the two layers. By computing the clustering coefficient for node i based on the contribution of nodes in layer 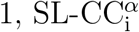, and nodes in layer 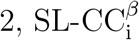, we calculate the ML-CC_i_ of node i as:

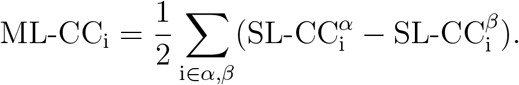

Then, the network multilayer clustering coefficient can be calculated as the average of the ML-CC_i_ over all nodes in the network.

#### Partial least squares regression analysis (PLS)

PLS performs a linear decomposition of the predictor and predicted variable matrix into latent variables (LVs) optimized so that the covariance between the resulting predictor and predicted matrix components (called factors and loadings respectively) is maximal [79]. We fit a PLS regression model for each predicted variable (cognitive tests, structural measures and prevalence of cardiovascular diseases) independently. The optimal component number for the decomposition was determined in each case by the Bayesian Information Criterion (BIC) based on the estimated degree of freedom of the PLS model [80]. In particular, predictor and predicted variable matrices were decomposed into 2-12 latent variables (LV), with the optimal number of LVs based on BIC scores being defined as follows: attention = 5, executive = 6, memory = 2, thickness = 12, visuospatial = 6, volumes = 5, WMH = 5, heart attack = 3, hypertension = The contribution of each variable to the prediction was quantified via Variable Importance in the Projection (VIP) scores, calculated as the sum of PLS-weights over LVs, weighted by the variance explained by each LV. Variables were defined as significant contributors to the prediction based on a VIP score of > 1 [81].

#### Genetic association analyses

We examined the association between 9 million high-quality imputed common variants (MAF > 1%, imputation INFO score > 0.8 and Hardy–Weinberg equilibrium P > 10^−10^) from UK Biobank and each derived phenotype using linear mixed-effects models as implemented in REGENIE [82, 83]. All traits were rank-based inverse normal transformed before the analysis, and adjusted for age at MRI, sex, age^2^, age*sex, age^2^*sex, body mass index (BMI), the first 10 principal components of ancestry, and genotyping array. For step 1, we used a subset of high-quality directly genotyped variants as described before [82]. We next performed a stringent physical LD clumping (PLINK parameters: –clump-p1 5e-8 –clump-r2 0.05 –clump-kb 1000, after excluding individuals with third-degree or closer relatives) [84, 85], followed by an approximate step-wise model selection in conditional and joint multiple-SNP analysis (COJO-GCTA) [86], with a window of 1 Mb and using 50,000 randomly selected unrelated Europeans (in-sample LD structure) as described before [82].

#### Colocalization

Colocalization was performed between independent genetic *loci* from COJO-GCTA and summary statistics of gene expression quantitative trait *loci* (eQTL) of 49 tissues in GTEx v8, BrainSeq, ROSMAP, Braineac2 and CommonMind from eQTL cataolgue release 4 [38–43]. All genes with at least one significant association (FDR adjusted p-value *<* 0.1) within a window of 1 Mb around each index variant were tested using coloc R package [87] with default priors, and H4 posterior probability (PP) > 0.8 was considered as a strong evidence that both traits share the same causal variant.

#### Statistical analysis

The statistical significance of the differences between men and women was assessed by performing nonparametric permutation tests with 10000 permutations, which were considered significant for a two-tailed test of the null hypothesis at p *<* 0.05. These results were adjusted for multiple comparisons by applying false discovery rate (FDR) corrections at q *<* 0.05 using the Benjamini–Hochberg procedure [88]. To detect sex differences over aging, we used linear regression models where all functional connectivity measures for individual subjects at each age were summarized by their average value due to the high overlap between brain measures across different ages. Then, they were included as the dependent variables in the linear regression models, whereas age, sex, age^2^, age*sex and age^2^*sex were included as independent variables. The best model for each measure (Supplementary Table S1) consisted of a combination of predictors that resulted in the minimum value of the Akaike information criterion (AIC) for that model. The significance of the overall model and the independent coefficients was evaluated by a F-test, which was considered significant at p *<* 0.05. To compare the performance of the different models, we used the Akaike information criterion (AIC) and the mean squared error of all models.

## Data Availability

The data used is part of UK Biobank cohort (https://www.ukbiobank.ac.uk/) and can be accessed by submitting an application to the UK Biobank research core.

