## Supplementary Appendix for "Sex differences in functional network topology over the course of aging in 37543 UK Biobank participants"

#### Supplementary information

Mite Mijalkov,[1,*] Dániel Veréb,[1] Oveis Jamialahmadi,[2] Anna Canal Garcia,[1] Emiliano Gomez Ruiz,[3] Didac Vidal-Piñeiro,[4] Stefano Romeo,[2] Giovanni Volpe,[3] and Joana B. Pereira[1,5,*]

[1]*Department of Neurobiology, Care Sciences and Society, Karolinska Institutet, Stockholm, Sweden*

[2]*Department of Molecular and Clinical Medicine, Goteborg University, Goteborg, Sweden*

[3]*Department of Physics, Goteborg University, Goteborg, Sweden*

[4]*Department of Psychology, University of Oslo, Oslo, Norway*

[5]*Memory Research Unit, Department of Clinical Sciences Malmö, Lund University, Lund, Sweden*

### I. SAMPLE CHARACTERISTICS

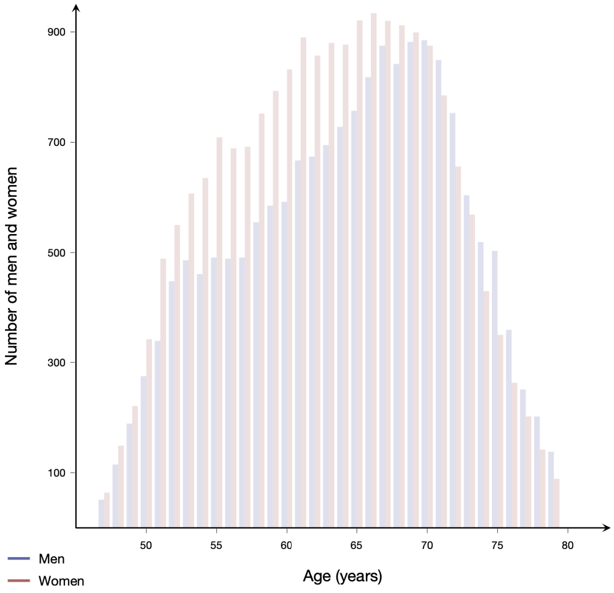


FIG. S1. **Sample characteristics.** Number of men (blue bars) and women (red bars) in the age range 47 - 79 years that were included in the current study.

#### A. Difference between men and women in different parameters.

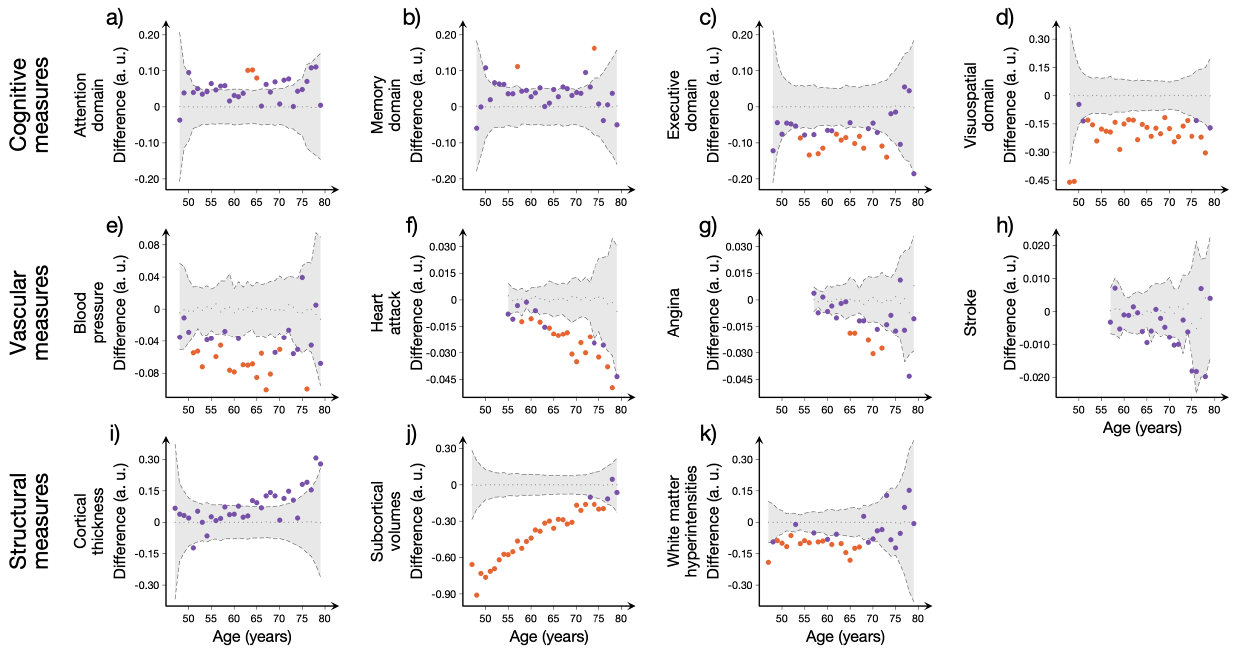


FIG. S2. **Differences between men and women in cognition, vascular and structural measures.** Plots showing the differences between men and women (calculated as women — men) in the a) attention, b) memory, c) executive and d) visuospatial cogntive domains. Differences between the prevalence of e) high blood pressure, f) heart attack, g) angina and h) stroke in men and women are also shown. Figure i-k show the observed sex differences in average cortical thickness, average subcortical volumes and white matter hyperintensities. The areas show the upper and lower bounds of the 95% confidence intervals (CI), and the differences in the corresponding measures between groups in blue circles as a function of individual's age. The differences are considered statistically significant if they fall outside the CIs. In particular, the orange circles show the differences that remained significant after applying a correction for multiple comparisons across the different age groups (FDR at $q < 0.05$).

#### II. CONNECTOGRAMS FOR MEN AND WOMEN AT DIFFERENT AGES.

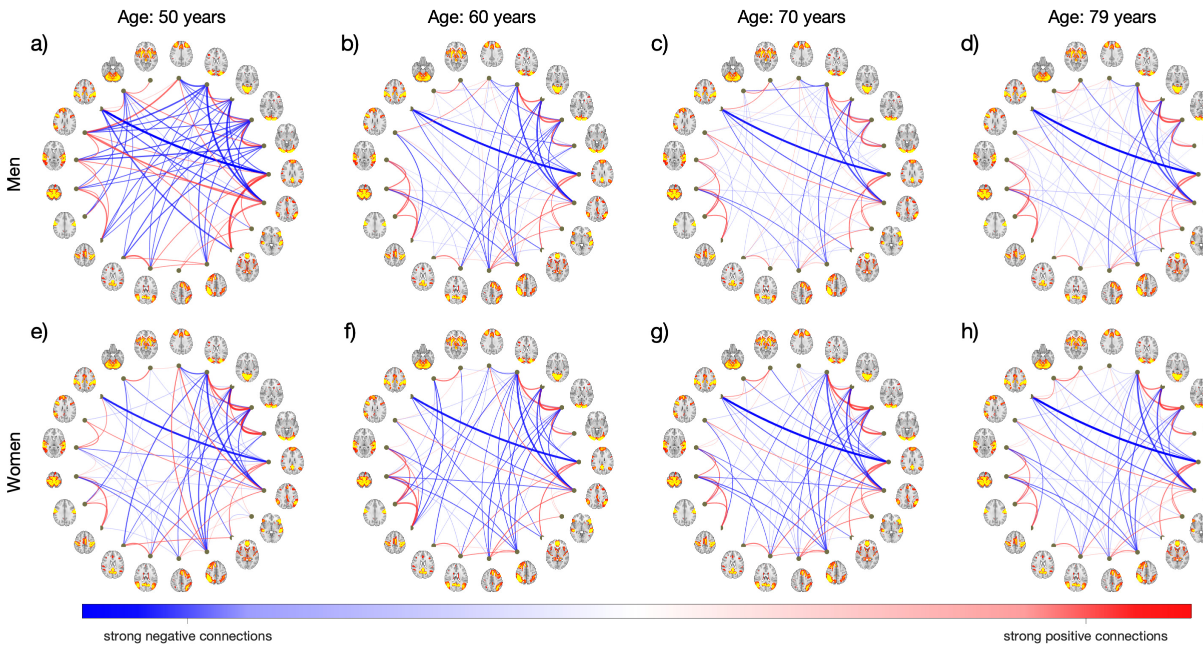


FIG. S3. **Connectogram-based representation of men's and women's average functional connectome at representative ages.** A representation of the men's and women's average functional connectivity networks at age of a-e) 50 years, b-f) 60 years, c-g) 70 years and d-h) 79 years of age. Thicker connections represent stronger functional connections; the positive and negative connections are shown in red and blue respectively. The 21 networks defined by group-ICA procedure represent the network nodes.

#### III. DIFFERENCES BETWEEN MEN AND WOMEN IN CONNECTIVITY, SINGLE LAYER AND MULTILAYER MEASURES.

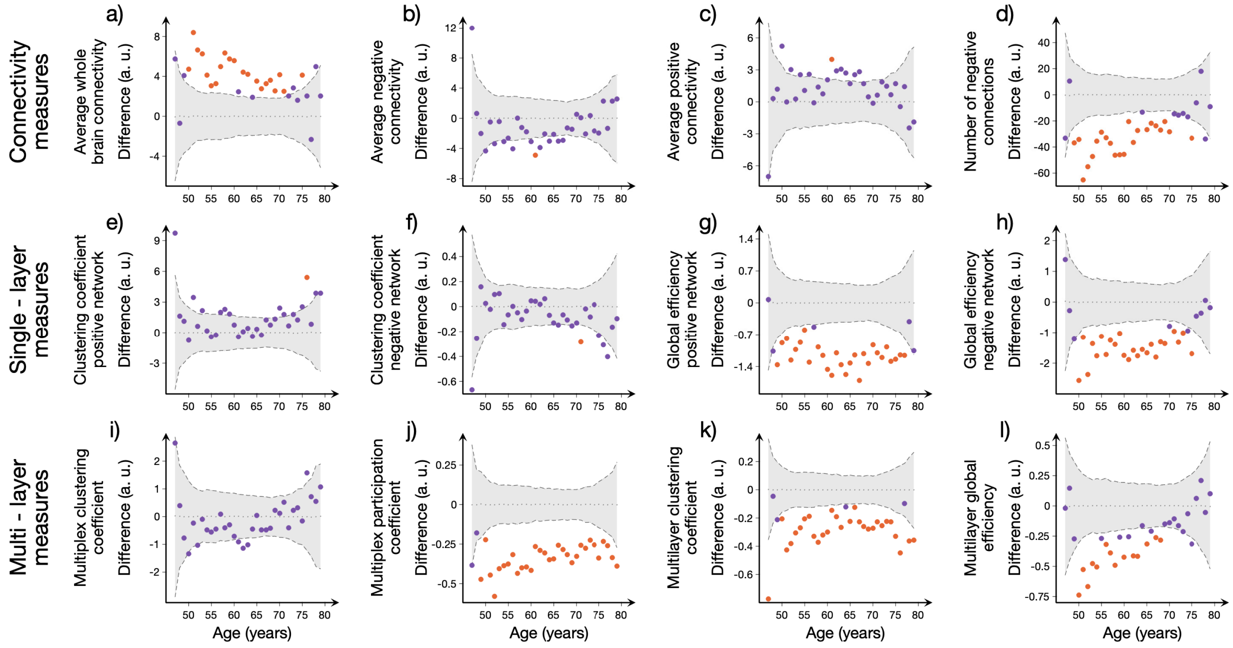


FIG. S4. **Differences between men and women in connectivity, single layer and multilayer measures.** Plots showing the differences between men and women (calculated as women - men) in a-d) simple connectivity measures: average whole-brain connectivity, average positive and negative connectivity and number of negative connections; e-f) single-layer topology measures: clustering coefficient and global efficiency of positive and negative networks; i-j) multiplex measures: clustering and participation coefficients and k-l) multilayer measures: clustering coefficient and global efficiency. The areas show the upper and lower bounds of the 95% confidence intervals (CI), and the differences in the corresponding measures between groups in blue circles as a function of individual's age. The differences are considered statistically significant if they fall outside the CIs. In particular, the orange circles show the differences that remained significant after applying a correction for multiple comparisons across the different age groups (FDR at $q < 0.05$)

#### IV. PREDICTION OF FUNCTIONAL CONNECTIVITY: MODEL SUMMARY

TABLE S1: Summary of the linear models used in the prediction of all measures of functional connectivity.

| | |
|---|---|
| Average connectivity $\sim 1 + Age^2 + Age * Sex$ | |
| Overall model | Coefficient (C) and p − values (pval) |
| $R^2 = 0.769$ | Age : C = 0.606; pval = 0.024 |
| AIC = 244.651 | $Age^2$ : C = −0.004; pval = 0.112 |
| MSE = 2.184 | Sex : C = −10.054; pval < 0.001 |
| F − stat = 55.2 | Age ∗ Sex : C = 0.101; pval = 0.010 |
| pval < 0.001 | |
| Average positive connectivity $\sim 1 + Age + Sex$ | |
| Overall model | Coefficient (C) and p − values (pval) |
| $R^2 = 0.403$ | Age : C = 0.163; pval < 0.001 |
| AIC = 281.229 | Sex : C = 1.138; pval = 0.023 |
| MSE = 3.947 | |
| F − stat = 22.9 | |
| pval =< 0.001 | |
| Average negative connectivity $\sim 1 + Age^2 + Sex$ | |
| Overall model | Coefficient (C) and p − values (pval) |
| $R^2 = 0.408$ | $Age^2$ : C = −0.001; pval < 0.001 |
| AIC = 236.801 | Sex : C = −1.149; pval = 0.002 |
| MSE = 2.013 | |
| F − stat = 23.4 | |
| pval < 0.001 | |
| Number of negative connections $\sim 1 + Age * Sex$ | |
| Overall model | Coefficient (C) and p − values (pval) |
| $R^2 = 0.778$ | Age : C = −0.932; pval < 0.001 |
| AIC = 501.106 | Sex : C = 83.806; pval < 0.001 |

Continued on next page

**TABLE S1 – continued from previous page**

| | |
|---|---|
| $MSE = 108.420$ | $Age * Sex: C = -0.895; pval < 0.001$ |
| $F - stat = 76.9$ | |
| $pval < 0.001$ | |
| **Clustering coefficient positive connections $\sim 1 + Age + Sex$** | |
| Overall model | Coefficient (C) and p – values (pval) |
| $R^2 = 0.459$ | $Age: C = 0.098; pval < 0.001$ |
| $AIC = 226.729$ | $Sex: C = -1.579; pval < 0.001$ |
| $MSE = 1.728$ | |
| $F - stat = 28.6$ | |
| $p - val < 0.001$ | |
| **Clustering coefficient negative connections $\sim 1 + Age + Sex$** | |
| Overall model | Coefficient (C) and p – values (pval) |
| $R^2 = 0.533$ | $Age: C = 0.012; pval < 0.001$ |
| $AIC = -96.294$ | $Sex: C = 0.096; pval = 0.001$ |
| $MSE = 0.013$ | |
| $F - stat = 38.1$ | |
| $pval < 0.001$ | |
| **Global efficiency positive connections $\sim 1 + Age^2 + Sex$** | |
| Overall model | Coefficient (C) and p – values (pval) |
| $R^2 = 0.924$ | $Age^2: C = 0.0005; pval < 0.001$ |
| $AIC = -1.051$ | $Sex: C = 1.089; pval < 0.001$ |
| $MSE = 0.055$ | |
| $F - stat = 396$ | |
| $pval < 0.001$ | |
| **Global efficiency negative connections $\sim 1 + Age^2 * Sex$** | |
| Overall model | Coefficient (C) and p – values (pval) |
| $R^2 = 0.649$ | $Age^2: C = 0.0002; pval = 0.006$ |

Continued on next page

**TABLE S1 – continued from previous page**

| | |
|---|---|
| $AIC = 88.603$ | $Sex: \ C = 1.750; \ pval < 0.001$ |
| $MSE = 0.209$ | $Age^2 * Sex: \ C = -0.0001; \ pval = 0.150$ |
| $F - stat = 41$ | |
| $pval < 0.001$ | |
| **Multiplex clustering coefficient $\sim 1 + Age + Age^2$** | |
| Overall model | Coefficient (C) and p – values (pval) |
| $R^2 = 0.096$ | $Age: \ C = -0.220; \ pval = 0.034$ |
| $AIC = 106.989$ | $Age^2: \ C = 0.002; \ pval = 0.048$ |
| $MSE = 0.282$ | |
| $F - stat = 4.45$ | |
| $pval = 0.016$ | |
| **Multiplex participation coefficient $\sim 1 + Age * Sex$** | |
| Overall model | Coefficient (C) and p – values (pval) |
| $R^2 = 0.891$ | $Age: \ C = -0.005; \ pval < 0.001$ |
| $AIC = -172.848$ | $Sex: \ C = 0.556; \ pval < 0.001$ |
| $MSE = 0.004$ | $Age * Sex: \ C = -0.003; \ pval < 0.036$ |
| $F - stat = 178$ | |
| $pval < 0.001$ | |
| **Multilayer clustering coefficient $\sim 1 + Age^2 + Sex$** | |
| Overall model | Coefficient (C) and p – values (pval) |
| $R^2 = 0.799$ | $Age^2: \ C = -0.0001; \ pval < 0.001$ |
| $AIC = -130.497$ | $Sex: \ C = 0.272; \ pval < 0.001$ |
| $MSE = 0.008$ | |
| $F - stat = 130$ | |
| $pval < 0.001$ | |
| **Multilayer global efficiency $\sim 1 + Age * Sex$** | |
| Overall model | Coefficient (C) and p – values (pval) |

Continued on next page

**TABLE S1 – continued from previous page**

| | |
|---|---|
| $R^2 = 0.795$ | $\text{Age}: \ C = -0.015; \ \text{pval} < 0.01$ |
| $\text{AIC} = -82.061$ | $\text{Sex}: \ C = 1.027; \ \text{pval} < 0.001$ |
| $\text{MSE} = 0.016$ | $\text{Age} * \text{Sex}: \ C = -0.012; \ \text{pval} < 0.001$ |
| $\text{F} - \text{stat} = 85.1$ | |
| $\text{pval} < 0.001$ | |

#### V. MULTILAYER MEASURES AS FUNCTION OF INTER-LAYER WEIGHT.

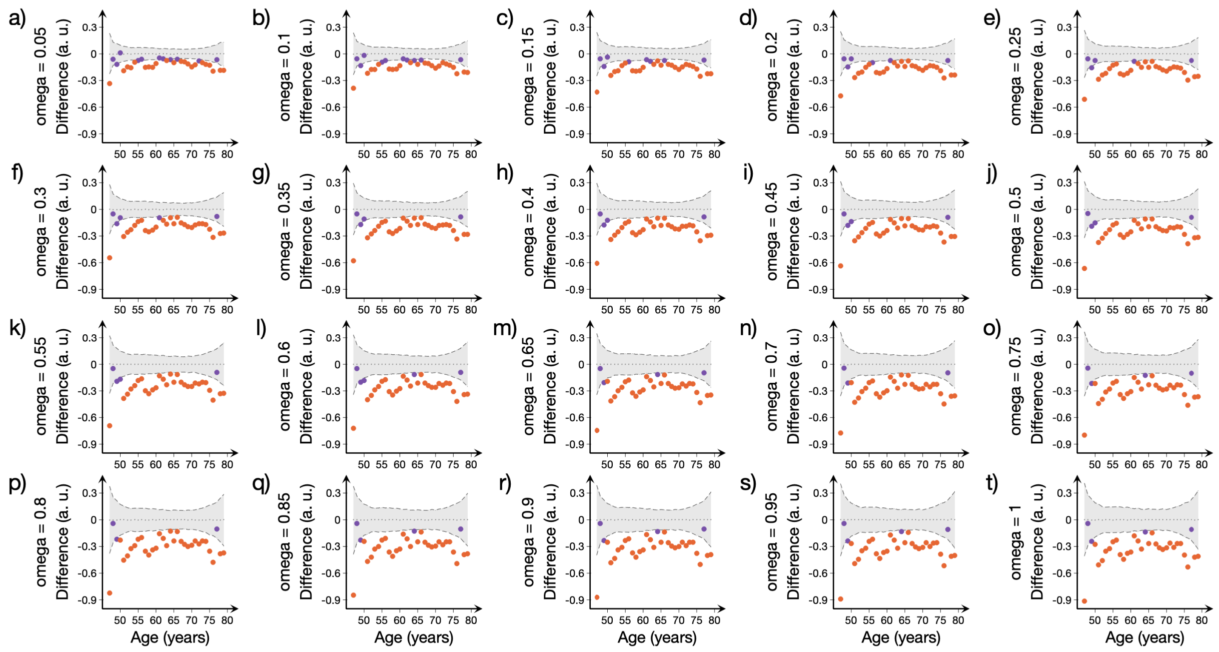


FIG. S5. **Differences between men and women in multilayer clustering**. Plots showing the differences between men and women (calculated as women — men) in the multilayer clustering for different values of inter-layer weight. The areas show the upper and lower bounds of the 95% confidence intervals (CI), and the differences in the corresponding measures between groups in blue circles as a function of individual's age. The differences are considered statistically significant if they fall outside the CIs. In particular, the orange circles show the differences that remained significant after applying a correction for multiple comparisons across the different age groups (FDR at $q < 0.05$).

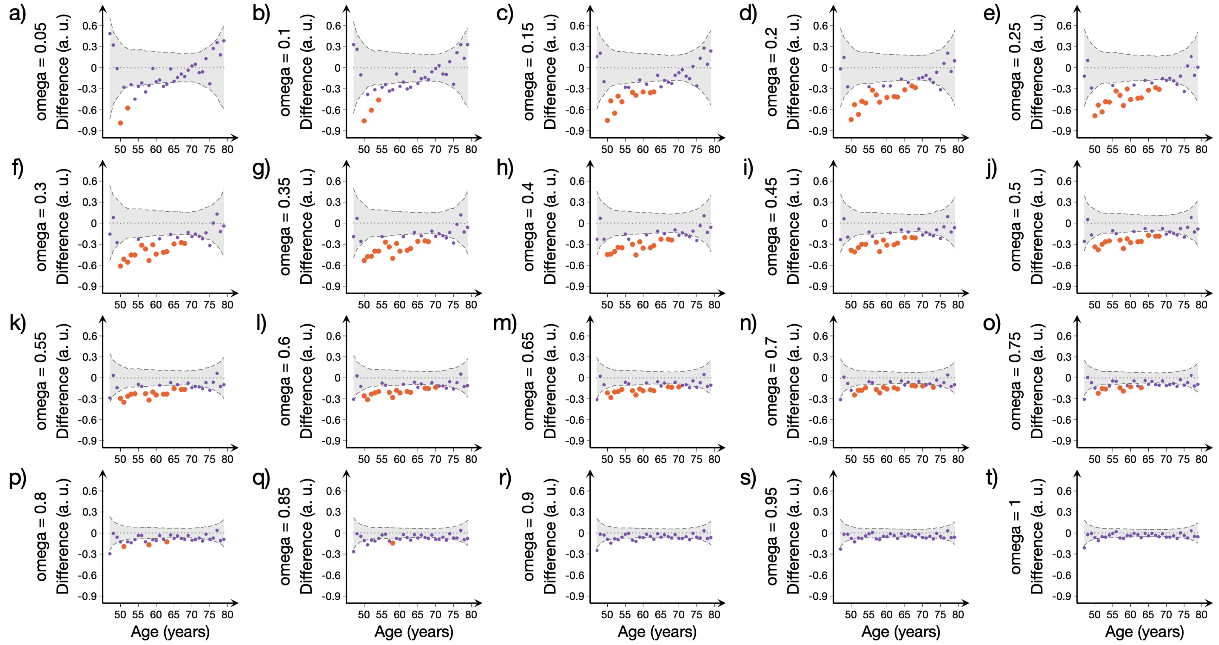


FIG. S6. **Differences between men and women in multilayer global efficiency**. Plots showing the differences between men and women (calculated as women — men) in the multilayer global efficiency for different values of inter-layer weight. The areas show the upper and lower bounds of the 95% confidence intervals (CI), and the differences in the corresponding measures between groups in blue circles as a function of individual's age. The differences are considered statistically significant if they fall outside the CIs. In particular, the orange circles show the differences that remained significant after applying a correction for multiple comparisons across the different age groups (FDR at $q < 0.05$).

#### VI. PARTIAL LEAST SQUARES REGRESSION ANALYSIS (PLS) DETAILS.

TABLE S2: **Relationship between executive domain test scores and connectivity, single-layer, multiplex and multilayer measures.** Details of the corresponding PLS analysis show the loadings in each latent variable, the VIP scores of each measure and the variance explained by the latent variables and the total model. Abbreviations: LV: latent variables; load.: loadings; Conn-Ave: Average connectivity; PosConn-Ave: Average positive connectivity; NegConn-Ave: Average negative connectivity; NegConn-No: Number of negative connections; SLpos-CC and SLneg-CC: Single layer clustering coefficient for networks of positive and negative connections; SLpos-Ge and SLneg-Ge: Single layer global efficiency for networks of positive and negative connections; MP-CC: Multiplex clustering coefficient; MP-Pt: Multiplex participation coefficient; ML-CC and ML-Ge: Multilayer clustering coefficient and global efficiency.

| **Executive domain** | | | | | | | |
|---|---|---|---|---|---|---|---|
| Variables | LV1 load. | LV2 load. | LV3 load. | LV4 load. | LV5 load. | LV6 load. | VIP scores |
| Age | 7,13 | 0,62 | 2,60 | 0,72 | 1,40 | -1,22 | 1,82 |
| Sex | 1,93 | 5,12 | -4,97 | 0,53 | -1,14 | 2,38 | 0,68 |
| Conn-ave | 3,42 | -4,94 | 4,79 | -0,43 | -1,04 | 1,55 | 0,54 |
| PosConn-Ave | 6,38 | -2,13 | -3,96 | -0,09 | 0,33 | 0,66 | 1,08 |
| NegConn-Ave | -6,01 | 1,54 | 4,40 | -1,02 | 0,75 | 1,46 | 1,06 |
| NegConn-No | -2,98 | 4,62 | -5,36 | -0,14 | 1,52 | -0,82 | 0,53 |
| SLpos-CC | 5,00 | -5,83 | 0,25 | -0,30 | 1,19 | 1,26 | 0,81 |
| SLneg-CC | 4,72 | 3,96 | 2,59 | -3,91 | 0,85 | -0,78 | 1,29 |
| SLpos-Ge | 6,77 | 2,93 | -2,42 | 1,13 | -0,64 | 0,01 | 1,57 |
| SLneg-Ge | 4,12 | 1,86 | -5,95 | 2,33 | -0,26 | 0,02 | 0,97 |
| MP-CC | 0,57 | -3,93 | -6,66 | 0,63 | 0,28 | -0,69 | 0,77 |
| MP-Pt | -0,65 | 5,33 | -5,49 | -0,11 | -0,35 | 0,61 | 0,35 |
| ML-CC | -3,68 | 5,63 | -3,98 | 0,48 | 0,03 | 0,81 | 0,59 |
| ML-Ge | -3,77 | 1,86 | -6,22 | 1,77 | 0,59 | -0,52 | 0,77 |
| **Variance explained by latent variables** | | | | | | | |

Continued on next page

**TABLE S2 – continued from previous page**

| | LV1 | LV2 | LV3 | LV4 | LV5 | LV6 | Total variance |
|---|---|---|---|---|---|---|---|
| Predictor matrix | 0,33 | 0,25 | 0,33 | 0,03 | 0,01 | 0,02 | 0,98 |
| Predicted variable | 0,72 | 0,08 | 0,02 | 0,08 | 0,02 | 0,00 | 0,92 |

TABLE S3: **Relationship between visuospatial domain test scores and connectivity, single-layer, multiplex and multilayer measures.** Details of the corresponding PLS analysis show the loadings in each latent variable, the VIP scores of each measure and the variance explained by the latent variables and the total model. Abbreviations are as Supplementary Table S2 and Fig. 4.

| **Visuospatial domain** | | | | | | | |
|---|---|---|---|---|---|---|---|
| Variables | LV1 load. | LV2 load. | LV3 load. | LV4 load. | LV5 load. | LV6 load. | VIP scores |
| Age | -6,49 | -4,08 | -0,98 | -0,84 | -1,52 | -0,10 | 1,76 |
| Sex | 3,03 | -5,88 | 3,59 | 2,24 | -0,55 | 0,36 | 0,84 |
| Conn-ave | -6,90 | 3,45 | -0,13 | 1,67 | -0,36 | 0,01 | 1,14 |
| PosConn-Ave | -4,12 | -2,93 | 5,75 | -1,36 | 0,52 | -0,85 | 0,77 |
| NegConn-Ave | 3,47 | 3,63 | -5,12 | 2,87 | -0,89 | -1,04 | 0,77 |
| NegConn-No | 6,66 | -3,51 | 1,04 | -1,40 | 0,51 | -0,90 | 1,08 |
| SLpos-CC | -6,46 | 1,62 | 3,60 | -1,38 | 0,50 | -1,21 | 1,05 |
| SLneg-CC | -3,25 | -5,33 | -3,28 | 2,14 | 2,64 | -1,29 | 0,99 |
| SLpos-Ge | -3,13 | -6,50 | 2,89 | 0,38 | -0,98 | 0,85 | 0,82 |
| SLneg-Ge | 0,09 | -5,33 | 5,36 | -1,60 | -0,47 | 1,01 | 0,25 |
| MP-CC | 0,68 | 0,72 | 6,31 | -4,29 | 0,65 | -1,16 | 0,40 |
| MP-Pt | 5,24 | -5,03 | 2,35 | 0,80 | -0,40 | -1,32 | 0,86 |
| ML-CC | 7,07 | -3,19 | 0,30 | 1,07 | -0,26 | -0,24 | 1,19 |
| ML-Ge | 6,68 | -1,14 | 2,82 | -2,45 | 0,51 | 0,67 | 1,18 |
| **Variance explained by latent variables** | | | | | | | |

Continued on next page

**TABLE S3 – continued from previous page**

| | LV1 | LV2 | LV3 | LV4 | LV5 | LV6 | Total variance |
|---|---|---|---|---|---|---|---|
| Predictor matrix | 0,40 | 0,27 | 0,21 | 0,06 | 0,02 | 0,01 | 0,98 |
| Predicted variable | 0,78 | 0,06 | 0,04 | 0,07 | 0,01 | 0,00 | 0,96 |

TABLE S4: **Relationship between prevalence of high blood pressure and connectivity, single-layer, multiplex and multilayer measures.** Details of the corresponding PLS analysis show the loadings in each latent variable, the VIP scores of each measure and the variance explained by the latent variables and the total model. Abbreviations are as Supplementary Table S2 and Fig. 4.

| **Blood pressure** | | | | | |
|---|---|---|---|---|---|
| Variables | LV1 load. | LV2 load. | LV3 load. | LV4 load. | VIP scores |
| Age | -0,58 | 1,12 | 1,35 | -0,73 | 0,24 |
| Sex | 7,56 | 2,21 | 0,12 | 0,88 | 1,78 |
| Conn-ave | -6,99 | 3,51 | 1,34 | 0,02 | 1,08 |
| PosConn-Ave | 1,46 | -0,30 | 7,53 | -1,69 | 0,40 |
| NegConn-Ave | -2,95 | 1,37 | -7,09 | 0,57 | 0,51 |
| NegConn-No | 7,00 | -3,28 | -0,52 | -0,65 | 1,06 |
| SLpos-CC | -3,69 | -0,84 | 6,97 | -0,14 | 0,64 |
| SLneg-CC | -3,69 | -0,84 | 6,97 | -0,14 | 0,64 |
| SLpos-Ge | 6,35 | 2,83 | 3,52 | -1,20 | 1,45 |
| SLneg-Ge | 6,08 | -1,34 | 4,94 | 0,43 | 1,02 |
| MP-CC | 1,34 | -3,35 | 6,44 | -2,78 | 0,62 |
| MP-Pt | 7,51 | 0,71 | -1,56 | -0,43 | 1,38 |
| ML-CC | 5,91 | 0,97 | -5,20 | 0,98 | 1,09 |
| ML-Ge | 3,52 | -5,54 | 3,91 | 1,96 | 0,79 |
| **Variance explained by latent variables** | | | | | |

Continued on next page

**TABLE S4 – continued from previous page**

| | LV1 | LV2 | LV3 | LV4 | Total variance |
|---|---|---|---|---|---|
| Predictor matrix | 0,41 | 0,09 | 0,36 | 0,02 | 0,89 |
| Predicted variable | 0,39 | 0,08 | 0,01 | 0,04 | 0,51 |

TABLE S5: **Relationship between prevalence of heart attack and connectivity, single-layer, multiplex and multilayer measures.** Details of the corresponding PLS analysis show the loadings in each latent variable, the VIP scores of each measure and the variance explained by the latent variables and the total model. Abbreviations are as Supplementary Table S2 and Fig. 4.

| **Heart attack** | | | | |
|---|---|---|---|---|
| Variables | LV1 load. | LV2 load. | LV3 load. | VIP scores |
| Age | -0,48 | -0,93 | 2,37 | 0,27 |
| Sex | 6,70 | 1,00 | 1,78 | 2,78 |
| Conn-ave | -1,66 | 1,19 | 5,98 | 0,38 |
| PosConn-Ave | 2,98 | -5,89 | 2,27 | 0,44 |
| NegConn-Ave | -2,89 | 5,96 | -2,26 | 0,48 |
| NegConn-No | 2,26 | -2,29 | -6,02 | 0,75 |
| SLpos-CC | 0,97 | -5,89 | 3,13 | 0,63 |
| SLneg-CC | 0,97 | -5,89 | 3,13 | 0,63 |
| SLpos-Ge | 4,75 | -3,54 | 2,37 | 1,08 |
| SLneg-Ge | 4,57 | -5,27 | 0,35 | 0,82 |
| MP-CC | 2,76 | -6,30 | 1,38 | 0,40 |
| MP-Pt | 4,54 | 1,55 | -3,74 | 1,30 |
| ML-CC | 0,38 | 5,68 | -4,08 | 0,67 |
| ML-Ge | 1,57 | -4,29 | -3,09 | 0,36 |
| **Variance explained by latent variables** | | | | |

Continued on next page

**TABLE S5 – continued from previous page**

| | LV1 | LV2 | LV3 | Total variance |
|---|---|---|---|---|
| Predictor matrix | 0,20 | 0,39 | 0,22 | 0,81 |
| Predicted variable | 0,48 | 0,09 | 0,04 | 0,61 |

TABLE S6: **Relationship between subcortical volumes and connectivity, single-layer, multiplex and multilayer measures.** Details of the corresponding PLS analysis show the loadings in each latent variable, the VIP scores of each measure and the variance explained by the latent variables and the total model. Abbreviations are as Supplementary Table S2 and Fig. 4.

| **Subcortical volumes** | | | | | | |
|---|---|---|---|---|---|---|
| Variables | LV1 load. | LV2 load. | LV3 load. | LV4 load. | LV5 load. | VIP scores |
| Age | -6,32 | -4,19 | 1,39 | -2,12 | -0,42 | 1,43 |
| Sex | 4,19 | -4,93 | 4,62 | 0,43 | -0,54 | 0,85 |
| Conn-Ave | -7,74 | 1,22 | -0,22 | 1,72 | -0,37 | 1,34 |
| PosConn-Ave | -3,71 | -0,29 | 6,58 | -2,51 | -0,03 | 0,63 |
| NegConn-Ave | 2,93 | 0,78 | -6,67 | 2,93 | -0,64 | 0,51 |
| NegConn-No | 7,65 | -1,07 | 0,82 | -1,88 | 0,22 | 1,32 |
| SLpos-CC | -6,28 | 3,11 | 3,18 | -1,62 | 0,99 | 1,02 |
| SLneg-CC | -3,12 | -6,98 | -0,58 | 0,56 | 2,06 | 0,82 |
| SLpos-Ge | -2,07 | -5,29 | 5,34 | -1,51 | -1,22 | 0,55 |
| SLneg-Ge | 1,97 | -2,64 | 6,79 | -2,47 | 0,65 | 0,34 |
| MP-CC | 1,69 | 4,62 | 5,05 | -3,54 | -0,34 | 0,51 |
| MP-Pt | 6,37 | -3,40 | 3,11 | -0,16 | -0,60 | 1,09 |
| ML-CC | 7,48 | -2,77 | 0,11 | 0,66 | -0,24 | 1,26 |
| ML-Ge | 7,42 | 1,33 | 1,62 | -1,35 | 1,61 | 1,38 |
| **Variance explained by latent variables** | | | | | | |
| | LV1 | LV2 | LV3 | LV4 | LV5 | Total variance |

Continued on next page

**TABLE S6 – continued from previous page**

| | | | | | | |
|---|---|---|---|---|---|---|
| Predictor matrix | 0,45 | 0,20 | 0,26 | 0,06 | 0,01 | 0,98 |
| Predicted variable | 0,84 | 0,03 | 0,01 | 0,04 | 0,01 | 0,93 |

TABLE S7: **Relationship between white matter hyperintensities and connectivity, single-layer, multiplex and multilayer measures.** Details of the corresponding PLS analysis show the loadings in each latent variable, the VIP scores of each measure and the variance explained by the latent variables and the total model. Abbreviations are as Supplementary Table S2 and Fig. 4.

| **White matter hyperintensities** | | | | | | |
|---|---|---|---|---|---|---|
| Variables | LV1 load. | LV2 load. | LV3 load. | LV4 load. | LV5 load. | VIP scores |
| Age | 7,48 | 1,62 | 1,74 | 1,27 | 0,29 | 1,72 |
| Sex | -0,75 | 7,33 | -2,48 | -1,50 | 0,88 | 0,54 |
| Conn-ave | 6,20 | -4,48 | 1,74 | -1,60 | 0,43 | 0,98 |
| PosConn-Ave | 6,06 | 1,03 | -5,01 | 0,59 | 0,06 | 1,07 |
| NegConn-Ave | -5,45 | -1,84 | 5,27 | -1,22 | 0,51 | 1,02 |
| NegConn-No | -5,92 | 4,57 | -2,26 | 1,54 | -0,38 | 0,92 |
| SLpos-CC | 5,91 | -4,20 | -3,00 | 0,38 | -1,18 | 0,89 |
| SLneg-CC | 4,01 | 4,71 | 4,58 | -0,19 | -1,89 | 1,21 |
| SLpos-Ge | 5,37 | 5,46 | -2,10 | -0,11 | 1,04 | 1,29 |
| SLneg-Ge | 1,48 | 5,31 | -5,69 | 0,38 | -0,79 | 0,51 |
| MP-CC | -0,75 | -1,16 | -7,20 | 2,86 | 0,54 | 0,54 |
| MP-Pt | -3,55 | 6,66 | -2,29 | -0,51 | 0,52 | 0,49 |
| ML-CC | -5,80 | 5,48 | -0,22 | -0,48 | 0,17 | 0,84 |
| ML-Ge | -6,34 | 2,51 | -3,84 | 0,90 | -1,47 | 1,14 |
| **Variance explained by latent variables** | | | | | | |
| | LV1 | LV2 | LV3 | LV4 | LV5 | Total variance |

Continued on next page

**TABLE S7 – continued from previous page**

| | | | | | | |
|---|---|---|---|---|---|---|
| Predictor matrix | 0,40 | 0,31 | 0,23 | 0,02 | 0,01 | 0,97 |
| Predicted variable | 0,78 | 0,07 | 0,02 | 0,07 | 0,01 | 0,95 |

#### VII. GENOME-WIDE ASSOCIATION STUDY (GWAS) RESULTS: QUANTILE-QUANTILE PLOTS.

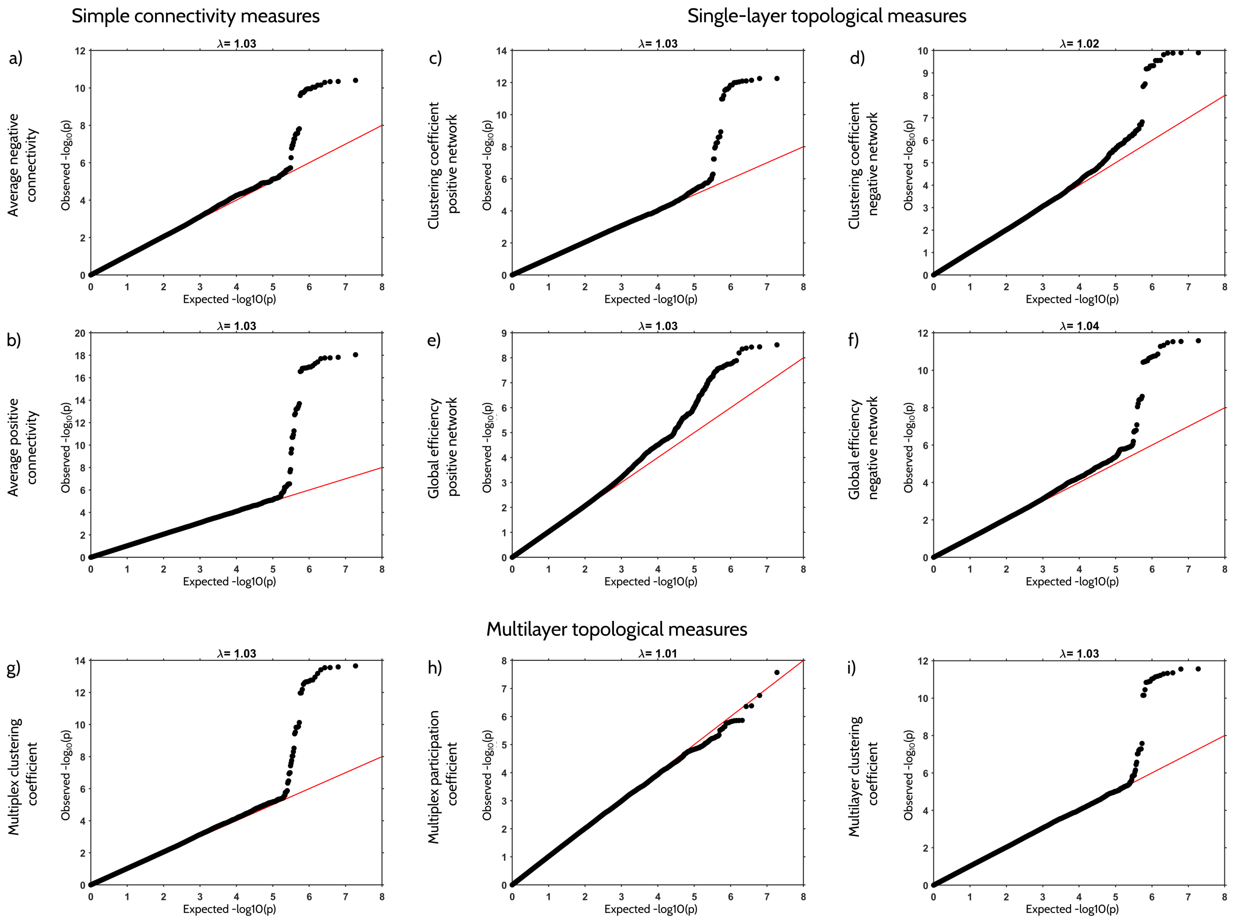


FIG. S7. **The quantile-quantile (QQ) plots for all functional connectivity measures that showed significant association with given genes**. QQ plots for all functional measures genomic inflation control (lambda) value. The red line shows the 95% confidence interval under the null hypothesis that there is no association among functional measures and SNPs. The black dots show the p-values of the complete study.

#### VIII. DENSITY OF THE POSITIVE AND NEGATIVE NETWORKS.

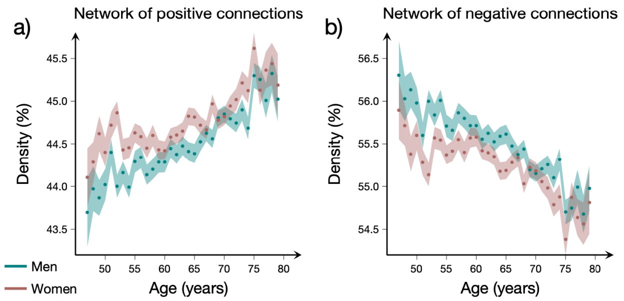


FIG. S8. **Density of the networks of positive and negative connections between men and women**. Plots showing the average density of the network of a) positive and b) negative connections for men (green) and women (red). The dots represent the mean density at a given age, while the shaded areas represent the standard error of the mean.

#### IX. ILLUSTRATION OF THE AREA UNDER THE CURVE (AUC) ANALYSIS.

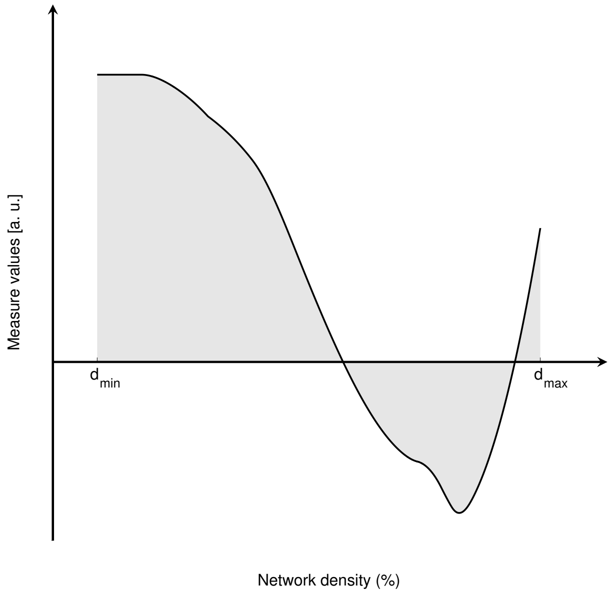


FIG. S9. **Illustration of the AUC analysis for the functional connectivity measures**. Each weighted connectivity network was binarized at a density range 6% to 33% in steps of 1%. We calculated each functional connectivity measure at all densities within this range and plotted the measure as a function of density (solid black line). For each measure, we integrated the total area under the curve (gray area). Calculated in this way, the AUC measure summarizes the behavior of the corresponding measure over the complete density range considered, and as such, it is less sensitive to the thresholding process.
