## Supplementary Table S8 for "Sex differences in functional network topology over the course of aging in 37543 UK Biobank participants"

Table S8. **Results of genome-wide association study of brain traits and colocalization.** Genome-wide significant (P < 5E-8) and independent (COJO-GCTA) variants for 9 brain traits have been shown. Colocalization was conducted using all variants withing a window of 1 Mb around each lead variant. Genes with a H4 posterior probability > 0.8 were considered to colocalize with the GWAS locus. Conn-Ave: Average connectivity; PosConn-Ave: Average positive connectivity; SLpos-CC and SLneg-CC: Single layer clustering coefficient for networks of positive and negative connections; SLpos-Ge and SLneg-Ge: Single layer global efficiency for networks of positive and negative connections; MP-CC: Multiplex clustering coefficient; MP-Pt: Multiplex participation coefficient; ML-CC: Multilayer clustering coefficient.

| FC measure | CHR | POS | SNP | Beta | SE | A1.freq | A2 | A1 | P | Nearest gene | eQTL.symbol | Max.H4.PP | Tissue(s) | Study |
|---|---|---|---|---|---|---|---|---|---|---|---|---|---|---|
| PosConn-Ave | 2 | 114081827 | rs62158169 | 0,083 | 0,009 | 0,786 | T | C | 8,9E-19 | PAX8 | IGKV1OR2-108 | 0,989 | esophagus (muscularis);lung;skin (suprapubic);skin | GTEx |
| | | | | | | | | | | | FOXD4L1 | 0,970 | thyroid | GTEx |
| | | | | | | | | | | | CBWD2 | 0,966 | adrenal gland | GTEx |
| MP-CC | 2 | 114081827 | rs62158169 | 0,071 | 0,009 | 0,786 | T | C | 2,2E-14 | PAX8 | IGKV1OR2-108 | 0,990 | esophagus (muscularis);lung;skin (suprapubic);skin | GTEx |
| | | | | | | | | | | | FOXD4L1 | 0,968 | thyroid | GTEx |
| | | | | | | | | | | | CBWD2 | 0,965 | adrenal gland | GTEx |
| SLpos-CC | 2 | 114103966 | rs56093896 | 0,068 | 0,009 | 0,788 | A | C | 5,6E-13 | PAX8 | IGKV1OR2-108 | 0,976 | esophagus (muscularis);lung;skin (suprapubic);skin | GTEx |
| | | | | | | | | | | | FOXD4L1 | 0,949 | thyroid | GTEx |
| | | | | | | | | | | | CBWD2 | 0,940 | adrenal gland | GTEx |
| SLneg-Ge | 2 | 114089551 | rs2863957 | 0,065 | 0,009 | 0,781 | A | C | 2,6E-12 | PAX8 | IGKV1OR2-108 | 0,992 | esophagus (muscularis);lung;skin (suprapubic);skin | GTEx |
| | | | | | | | | | | | FOXD4L1 | 0,976 | thyroid | GTEx |
| | | | | | | | | | | | CBWD2 | 0,967 | adrenal gland | GTEx |
| ML-CC | 2 | 114106139 | rs62158211 | -0,066 | 0,009 | 0,788 | T | G | 2,8E-12 | PAX8 | IGKV1OR2-108 | 0,976 | esophagus (muscularis);lung;skin (suprapubic);skin | GTEx |
| | | | | | | | | | | | FOXD4L1 | 0,943 | thyroid | GTEx |
| | | | | | | | | | | | CBWD2 | 0,936 | adrenal gland | GTEx |
| NegConn-Ave | 2 | 114081827 | rs62158169 | -0,062 | 0,009 | 0,786 | T | C | 4,0E-11 | PAX8 | IGKV1OR2-108 | 0,988 | esophagus (muscularis);lung;skin (suprapubic);skin | GTEx |
| | | | | | | | | | | | FOXD4L1 | 0,978 | thyroid | GTEx |
| | | | | | | | | | | | CBWD2 | 0,959 | adrenal gland | GTEx |
| SLneg-CC | 2 | 114110568 | rs62158213 | -0,060 | 0,009 | 0,789 | A | G | 1,3E-10 | PAX8 | IGKV1OR2-108 | 0,978 | esophagus (muscularis);lung;skin (suprapubic);skin | GTEx |
| | | | | | | | | | | | FOXD4L1 | 0,927 | thyroid | GTEx |
| | | | | | | | | | | | CBWD2 | 0,929 | adrenal gland | GTEx |
| SLpos-Ge | 2 | 114085785 | rs7556815 | 0,054 | 0,009 | 0,782 | A | G | 3,0E-09 | PAX8 | IGKV1OR2-108 | 0,993 | esophagus (muscularis);lung;skin (suprapubic);skin | GTEx |
| | | | | | | | | | | | FOXD4L1 | 0,977 | thyroid | GTEx |
| | | | | | | | | | | | CBWD2 | 0,967 | adrenal gland | GTEx |
| SLpos-Ge | 10 | 134300091 | rs4309079 | 0,043 | 0,008 | 0,484 | T | C | 1,8E-08 | INPP5A | INPP5A | 0,878 | brain (DLPFC) | CommonMind |
| MP-Pt | 13 | 105359436 | rs9514306 | 0,085 | 0,015 | 0,932 | C | T | 2,7E-08 | DAOA | - | - | - | - |
| MP-CC | 10 | 2002410 | rs2152237 | 0,173 | 0,031 | 0,985 | A | G | 3,7E-08 | ADARB2 | - | - | - | - |
